# Understanding the impact of antenatal care policies in Georgia (USA) and Scotland (UK): A textual synthesis

**DOI:** 10.1101/2024.03.21.24304668

**Authors:** J. Shim, V. Burnett, M. Vernon, F. Work, I. Uwaifo, C. Ray, P. Reddi

## Abstract

**Objectives:** This study aims to (1) understand the role of policy in maternal health outcomes, and (2) establish any differences or similarities between health systems, providing benchmarks for future maternal and infant care policies in Georgia and Scotland.

**Methods:** Guided by JBI methodology, a textual review of policies and public health interventions that have influenced the antenatal care process in both health systems was conducted. Inclusion criteria for this review were classified using the “PCC” mnemonic: Population-Pregnant women and mothers; Concept-Policies and strategies that supports prenatal and maternal health; and Context-Relevant to Scotland and Georgia. Published primary and secondary research, and grey literature (guidelines, reports, and legislation from authoritative sources) were included.

**Results:** Overall, 60 sources contributed to the report on maternal health system topics. Findings of the textual synthesis presented a regionalized system of maternity care led by physician-provided care models in Georgia compared to the nationalized health system in Scotland with an extended scope for midwife-led care models. On a secondary, organizational level, Scotland also widely operates on protocolized, standardized care informed by clinical guidelines such as NICE. The Georgia health systems also follow national guidelines for care, but extent of standardization may vary based on a mixed system of private and public insurance coverage.

**Discussion/Conclusion:** This is the first study to comprehensively examine maternal health policies in the distinct contexts of Georgia and Scotland, shedding light on the diverse approaches within their respective healthcare systems. These observed variations stem from historical, cultural, and policy contexts unique to each region. As the United States continue to prioritize maternal and child health through public health initiatives, our findings feature crucial considerations for maternal antenatal care policies. Specifically, there is a discernible need to increase access to antenatal care and invest in the maternity care provider workforce, revealing opportunities for targeted improvements in support of maternal health.

## Introduction

Maternal and infant health disparities are a pressing global concern, with the United States (US) experiencing the highest maternal and infant mortality rates among high-income nations.^1,2^ This rising trend in mortality rates has brought maternal and infant health to the forefront of many recent public health and research initiatives within the US.^3^

Georgia has the second highest maternal mortality rate and the twelfth highest infant mortality rate in the US.^4,5^ Similar to the rest of the US, research has indicated Black mothers in Georgia are more likely to die from pregnancy, especially in rural areas.^4^ Understanding the factors affecting maternal and infant health in these underserved regions is crucial in addressing healthcare disparities in the US.^6^

In comparison, while Georgia had a maternal mortality rate of 50.8 maternal deaths per 100,000 births in 2019, Scotland (UK) reported a lower rate of 10.9 maternal deaths per 100,000 births between 2017-2019.^7,8^ In 2021, 1,205 women died of maternal causes in the US compared to 861 women in 2020 and 754 women in 2019.^3^ Both rural regions in Scotland and Georgia face similar healthcare challenges, including a need for physicians and limited access to care.^9^ This shared context offers an important point of comparison between the healthcare systems that operate within these regions. Despite both countries being high-income nations with high rankings on the Human Development Index (HDI),^10^ they offer distinct healthcare systems and policies for comparison.

The US, with its decentralized and predominantly privatized healthcare system, faces unique challenges in ensuring equitable access to maternal care across diverse regions.^11^ In contrast, Scotland’s publicly funded and centralized healthcare system strives to provide comprehensive healthcare coverage, albeit with its own set of challenges.^12^ Understanding these variations in healthcare systems, their infrastructure, and policies in the context of maternal and infant health, may elucidate the factors contributing to disparities in preterm birth rates and inform discussions on maternal care quality and policy reforms in these regions. This understanding is not only valuable for the nations involved but also contributes to global efforts to reduce persistent inequalities in health, attainment, and life expectancies, especially in marginalized communities.^13,14^

Therefore, our study aims to (1) understand the role of policy in maternal health outcomes, and (2) establish any differences or similarities between health systems, providing benchmarks for future maternal and infant care policies in Georgia and Scotland.

## Methods

### Study Design

A comparative approach was conducted through a textual synthesis of published literature on the health systems and policies influencing maternal health in Georgia and Scotland. We recognized that health systems are complex and multifaceted, influenced by a multitude of variables including government policies, law, national infrastructure, socioeconomic conditions, cultural norms, and the interactions of various stakeholders – patients, providers, payors, and policymakers.^15^ Therefore, the intention of the textual review was to provide a nuanced understanding of the policy landscapes of each region to encourage any cross-regional learning.

### Textual Review of Maternal Health Literature

The textual review was conducted in accordance with JBI methodology.^16^

#### Inclusion criteria

Inclusion criteria for this review were classified using the “PICo” (Population, Phenomenon of interest, Context) mnemonic: Population: Pregnant and birthing women; Phenomenon of Interest: Policy strategies to reduce maternal morbidity and mortality; and Context: Health setting in the UK and the US.^16^

#### Type of sources

Expert opinions, consensus, current discourse, comments, assumptions, or assertions that appear in various formats including journals, magazines, newspapers, blogs, internet sites, monographs and reports were largely drawn for this analysis. We reviewed grey literature that contains policy- and research-relevant information (e.g., clinical practice guidelines, national reports, program evaluation studies, and legislation) from authoritative sources that are widely accessible.^17^

#### Search strategy

The search comprised of three steps; Firstly, a limited search of MEDLINE and CINAHL using initial keywords was conducted to develop a full search strategy. Secondly, the full search strategy was adapted to each database and applied systematically to: MEDLINE, CINAHL, AMED, EMBase, and Cochrane Library (see Appendix 1). Finally, the third step involved conducting a search of grey and unpublished literature in the Maternity and Infant Care database, Open Grey, MedNar, The New York Academy Grey Literature Report, Ethos, CORE, and Google Scholar using modified search terms for maternal health-related policies. We also searched the US Congress online legislative database and Georgia online legislative database for national and Georgia state-level proposed legislation and legislation passed between 2000 and 2020 addressing maternal health. No limit was placed on language, but all of the research studies published were in English.

#### Study selection

Following the search, all identified sources were collated and uploaded into EndNote and duplicates were removed. Sources were then imported to Covidence (Melbourne, Australia) for two-level screening. Firstly, titles and abstracts were screened independently by two reviewers (PR and VB) with conflicts identified by the management software and resolved by a third reviewer (JS and FW). Secondly, full-text copies of all sources included at the title and abstract screening stage were screened using the same processes.

#### Quality assessment

Due to the nature of grey literature, which was largely descriptive and based on expert opinions, it was not appropriate to critically appraise the evidence for this textual synthesis.

#### Data extraction and synthesis

A data extraction tool was developed for this review to extract relevant information about the study and key findings. Data that were extracted were synthesized with the use of tabulation and graphs and presented alongside an accompanying narrative. The synthesis was focused on data relating to similarities and differences between the countries.

## Results

### Findings from the textual synthesis

The initial search identified 846 articles in the databases, supplemented by a further 253 studies from the grey literature (websites and expert sources). 60 reports were included following full-text screening. Figure 1 presents the study selection process and the main reasons for exclusion.

**Figure 1.**
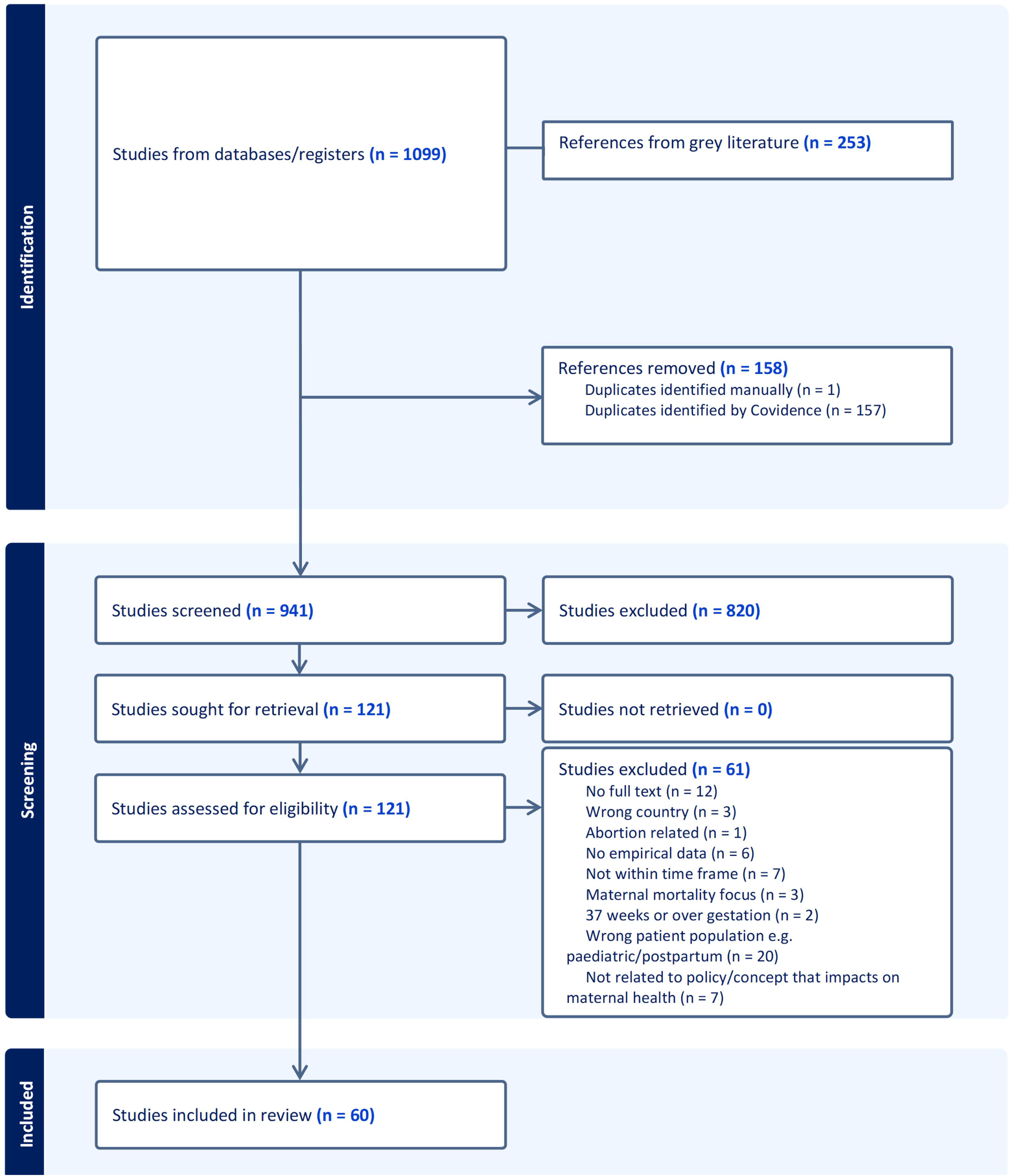
PRISMA flowchart of included studies.

A wide variety of different journals and policies contributed to the report on maternal health system topics, including the *Journal of Policy Analysis and Management*, *Maternal and Child Health Journal*, and the *World Health Organization*. Of the 60 included reports, 42 were related to Scottish policies while 18 were specific to Georgia. The majority of the studies were widely accessible public reports with a descriptive component to their design. 27 observational and experimental studies examined the contents of policy or its impact, and 7 were reviews of the literature. Table 1 presents a summary of the characteristics of the included studies.

**Table 1.**
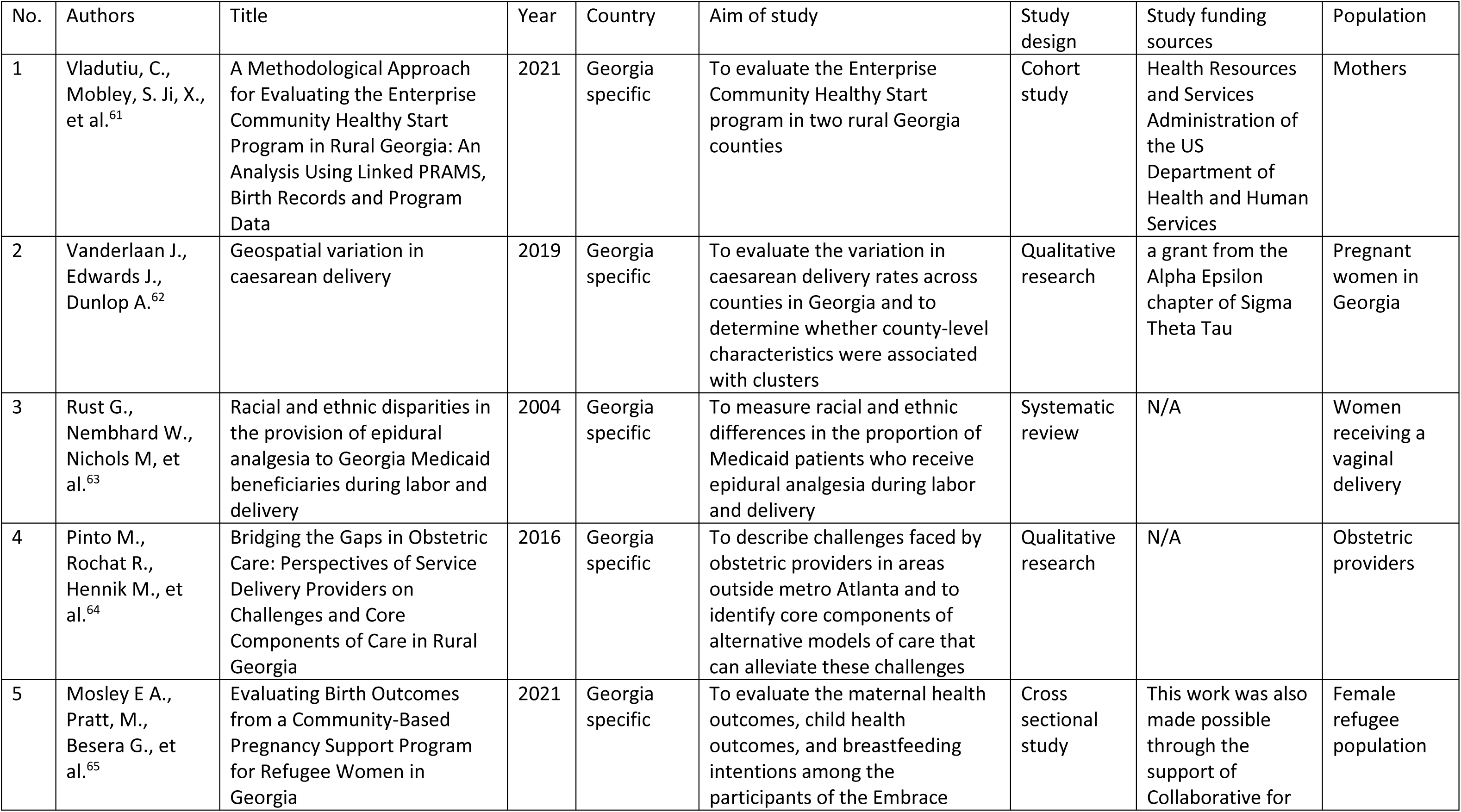
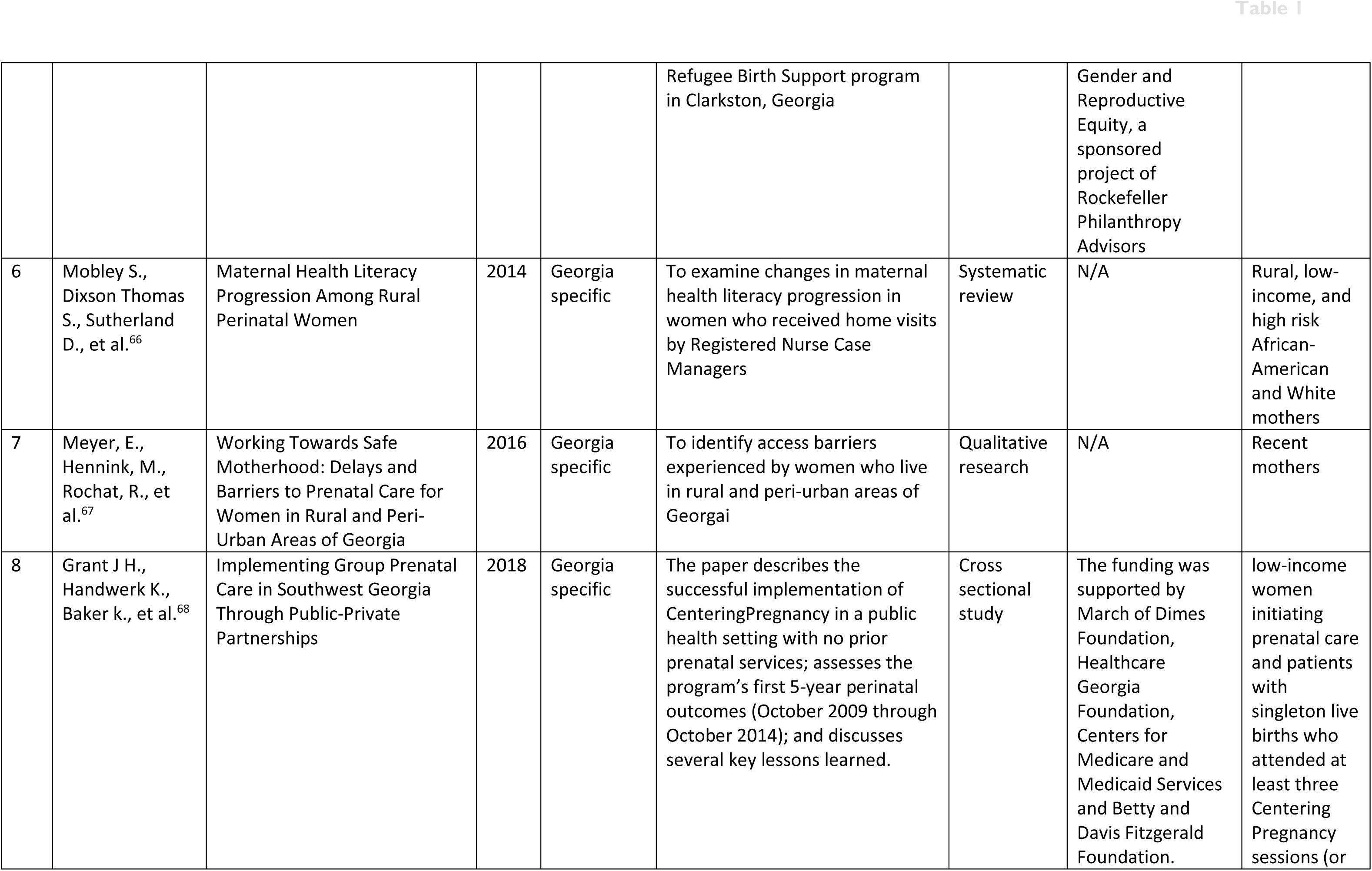
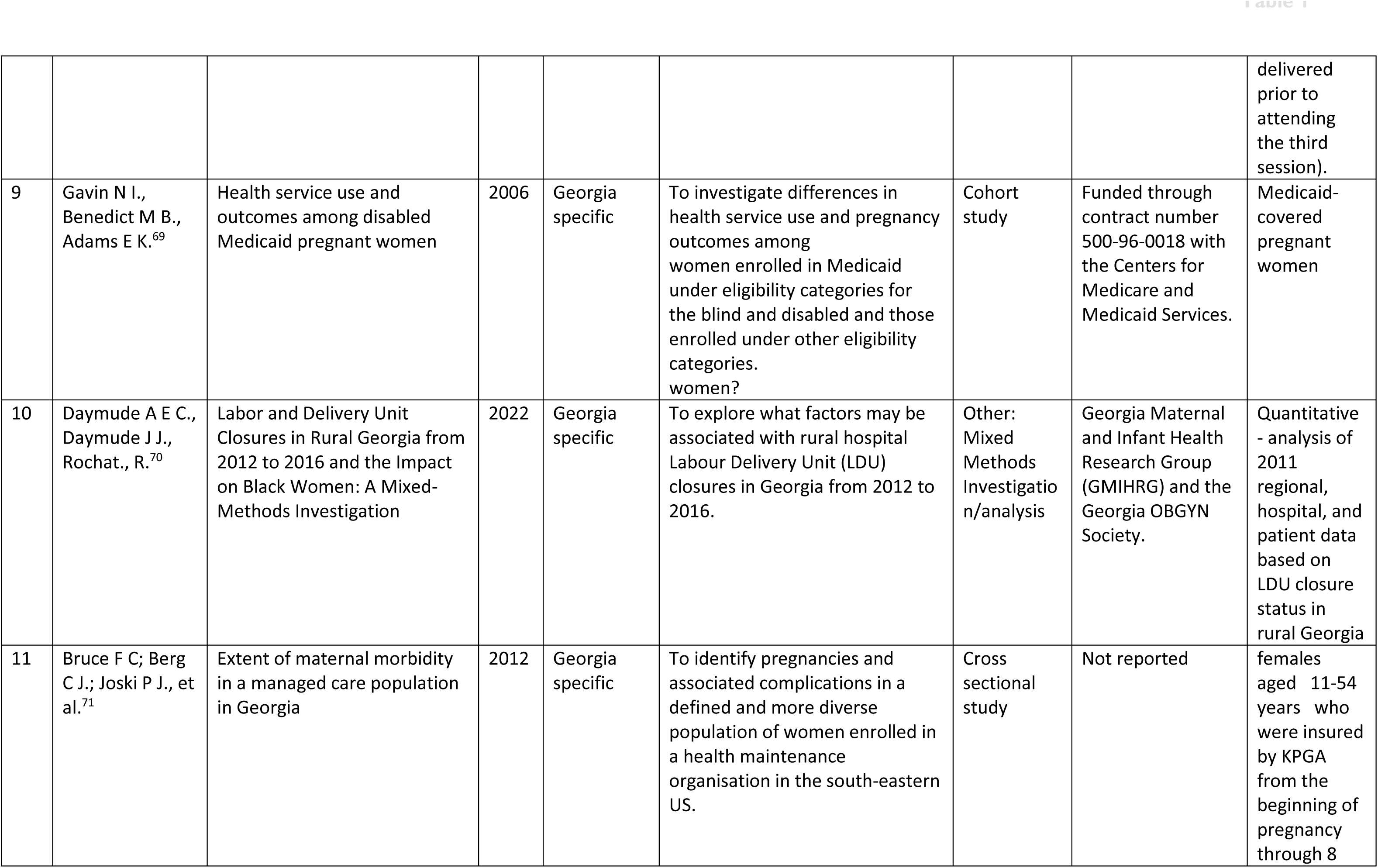
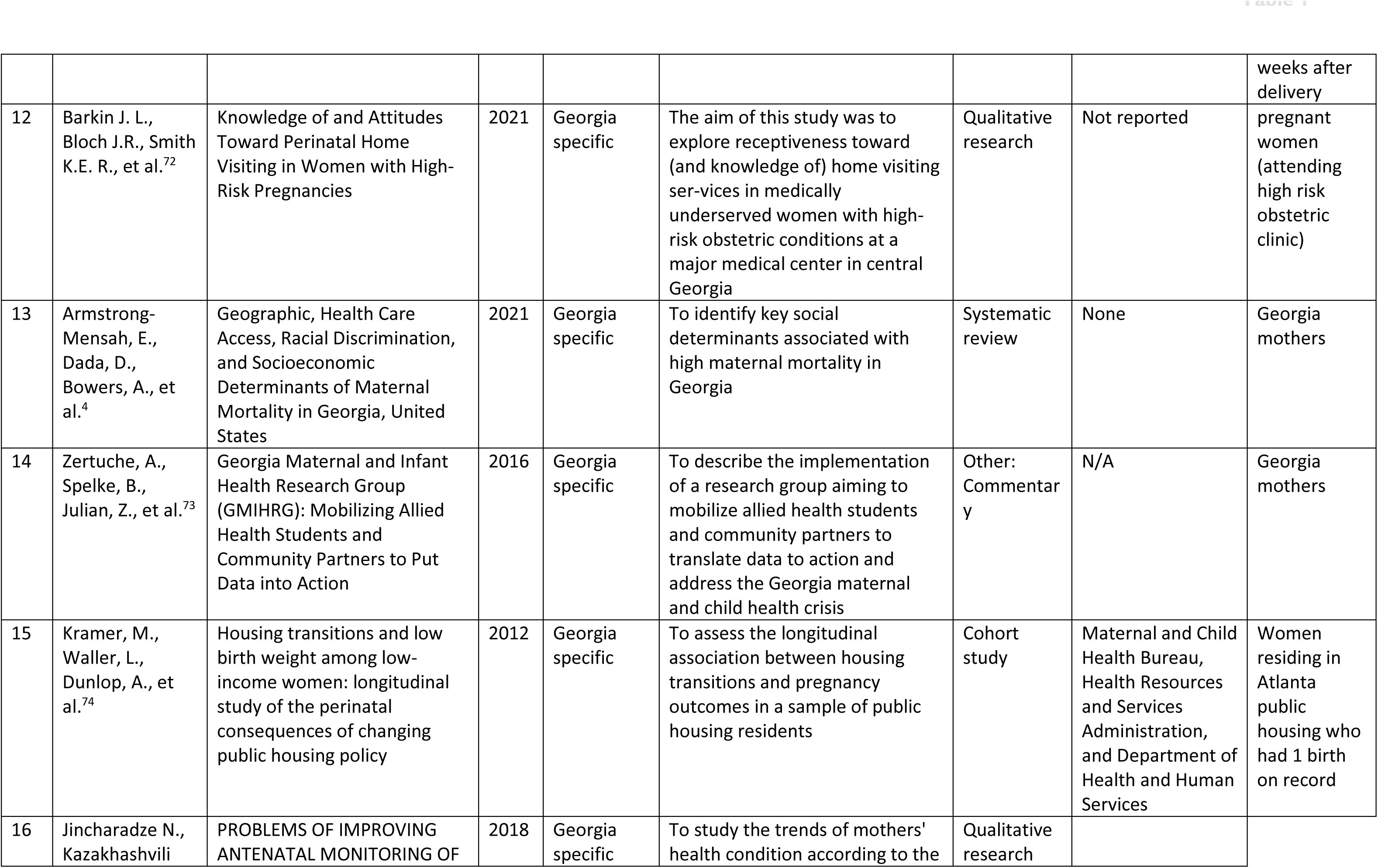
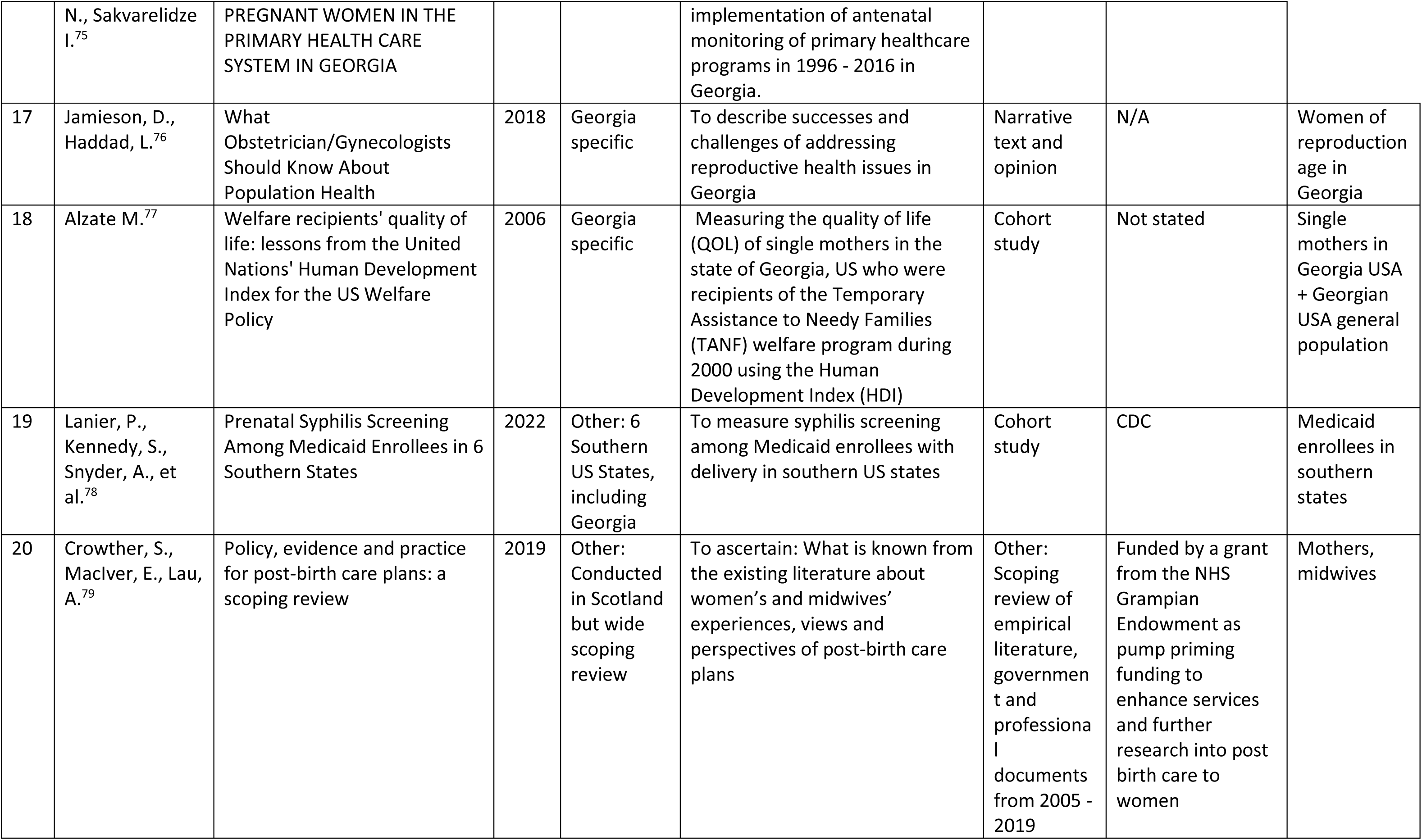
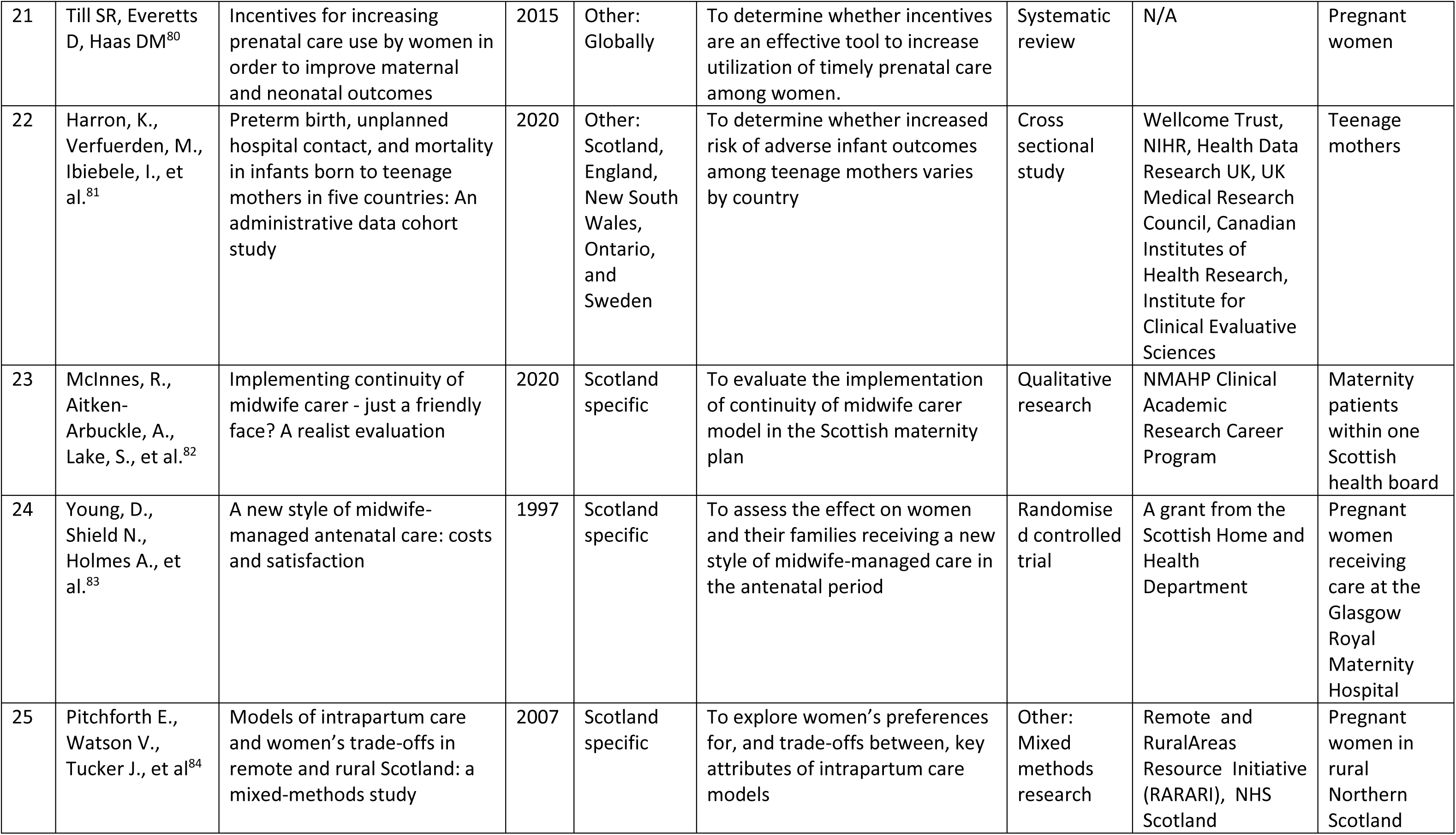
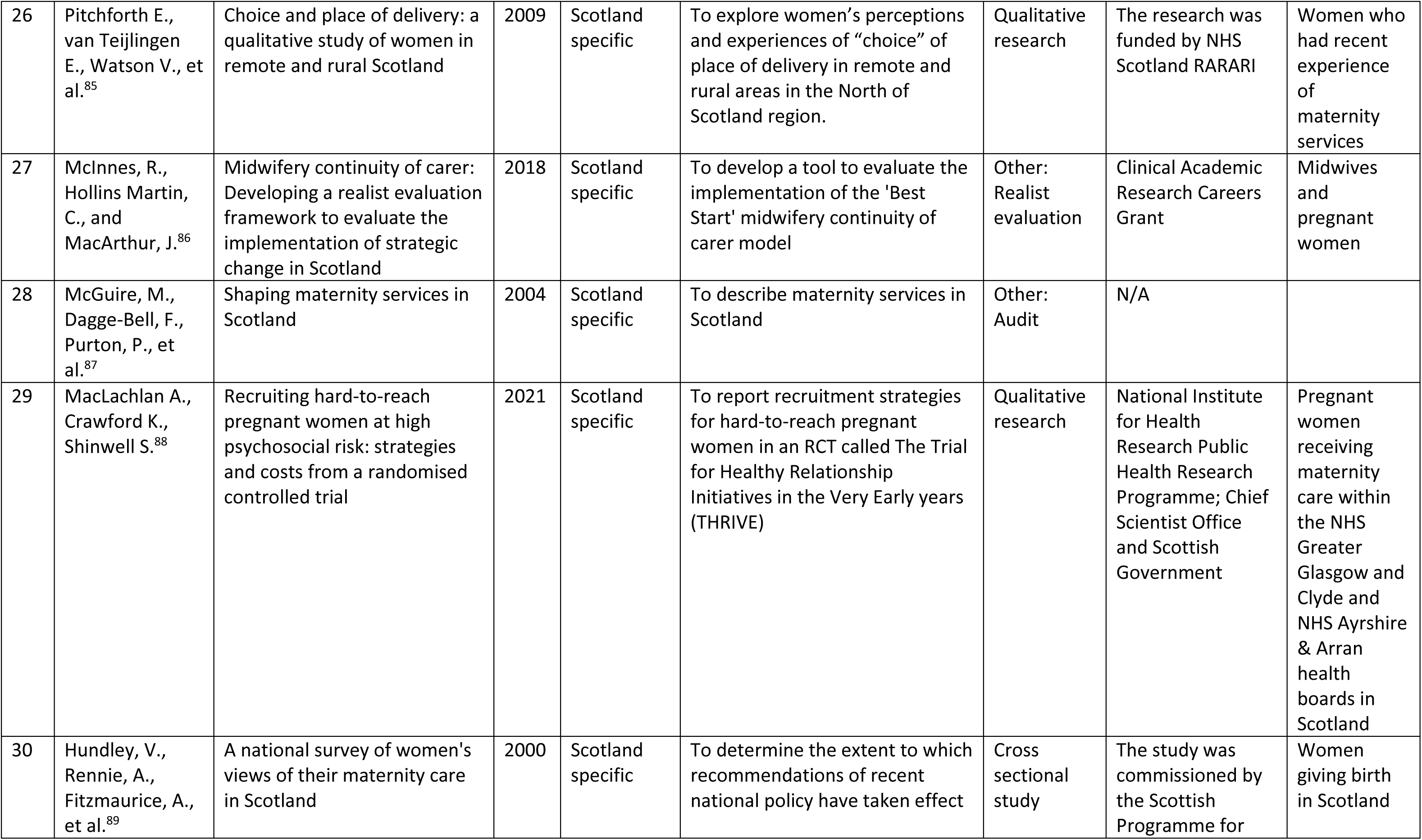
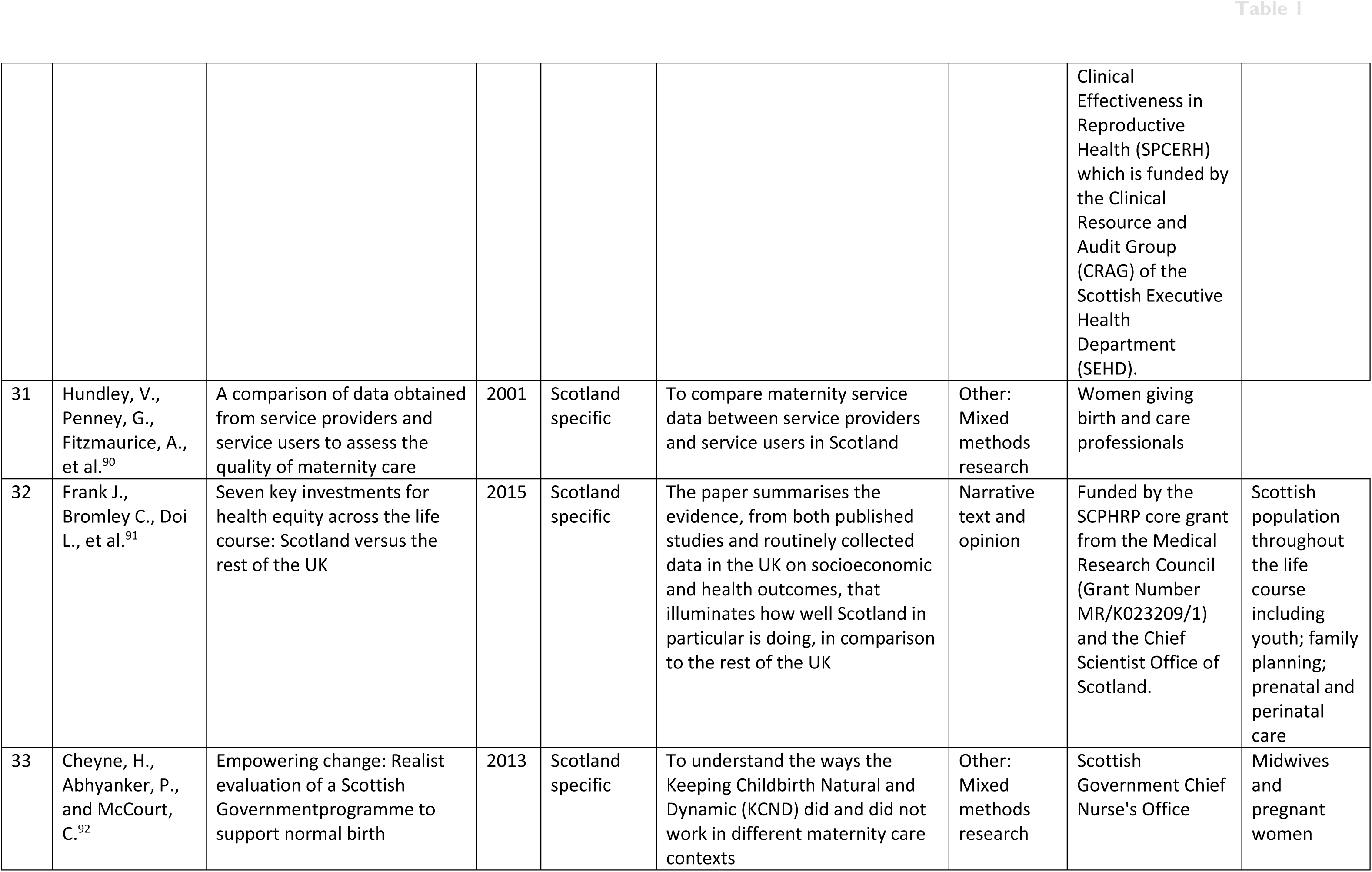
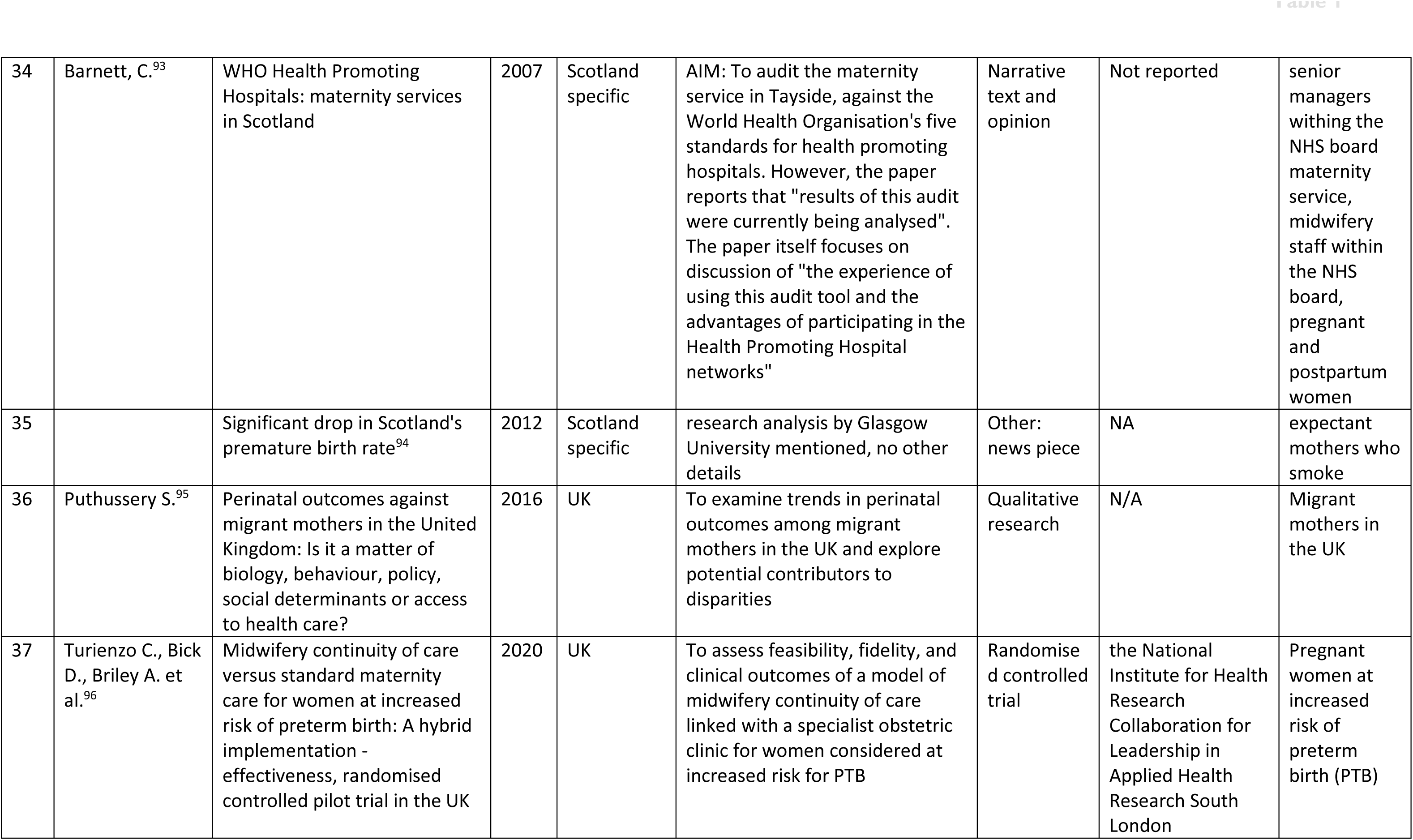
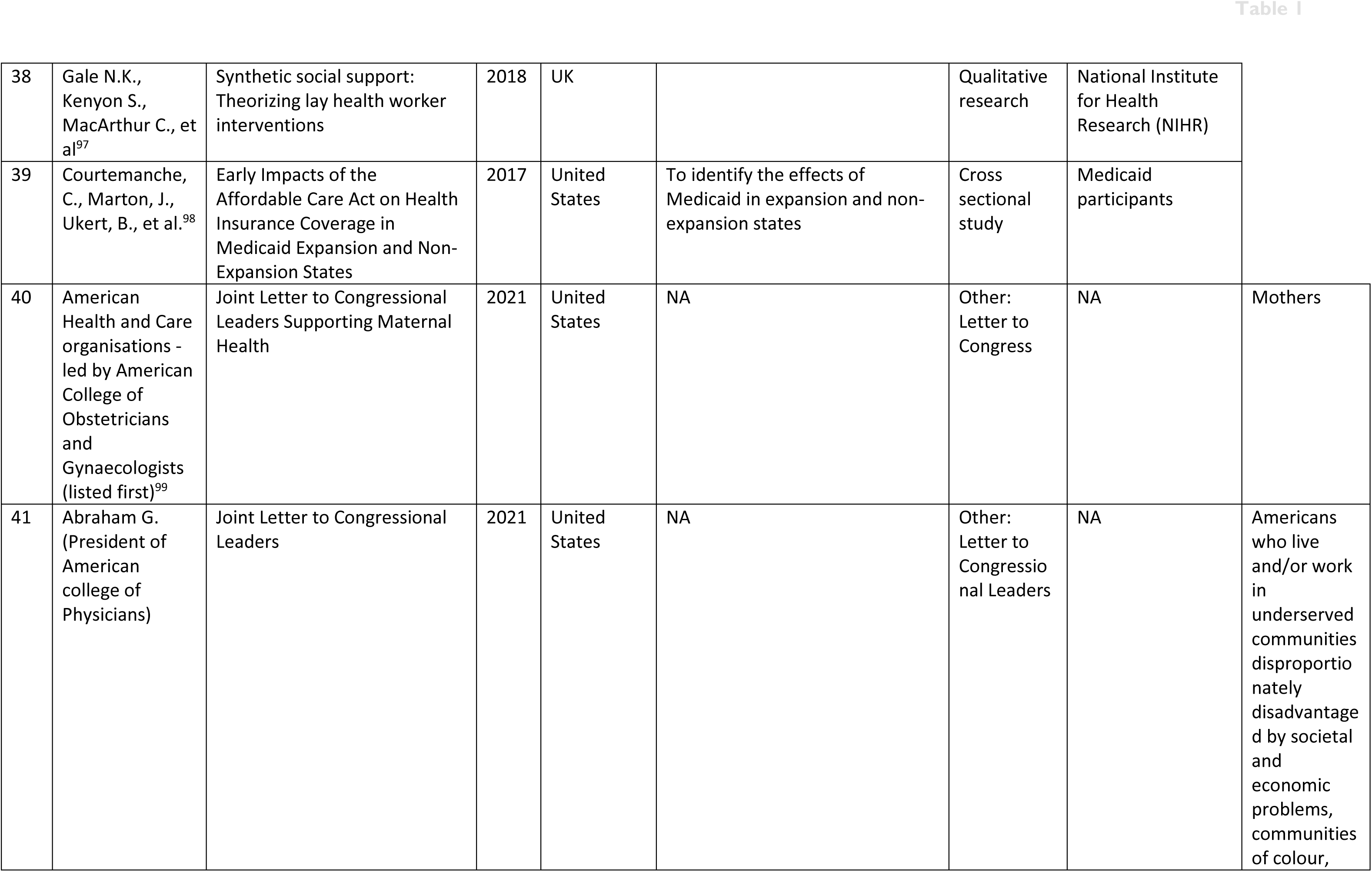
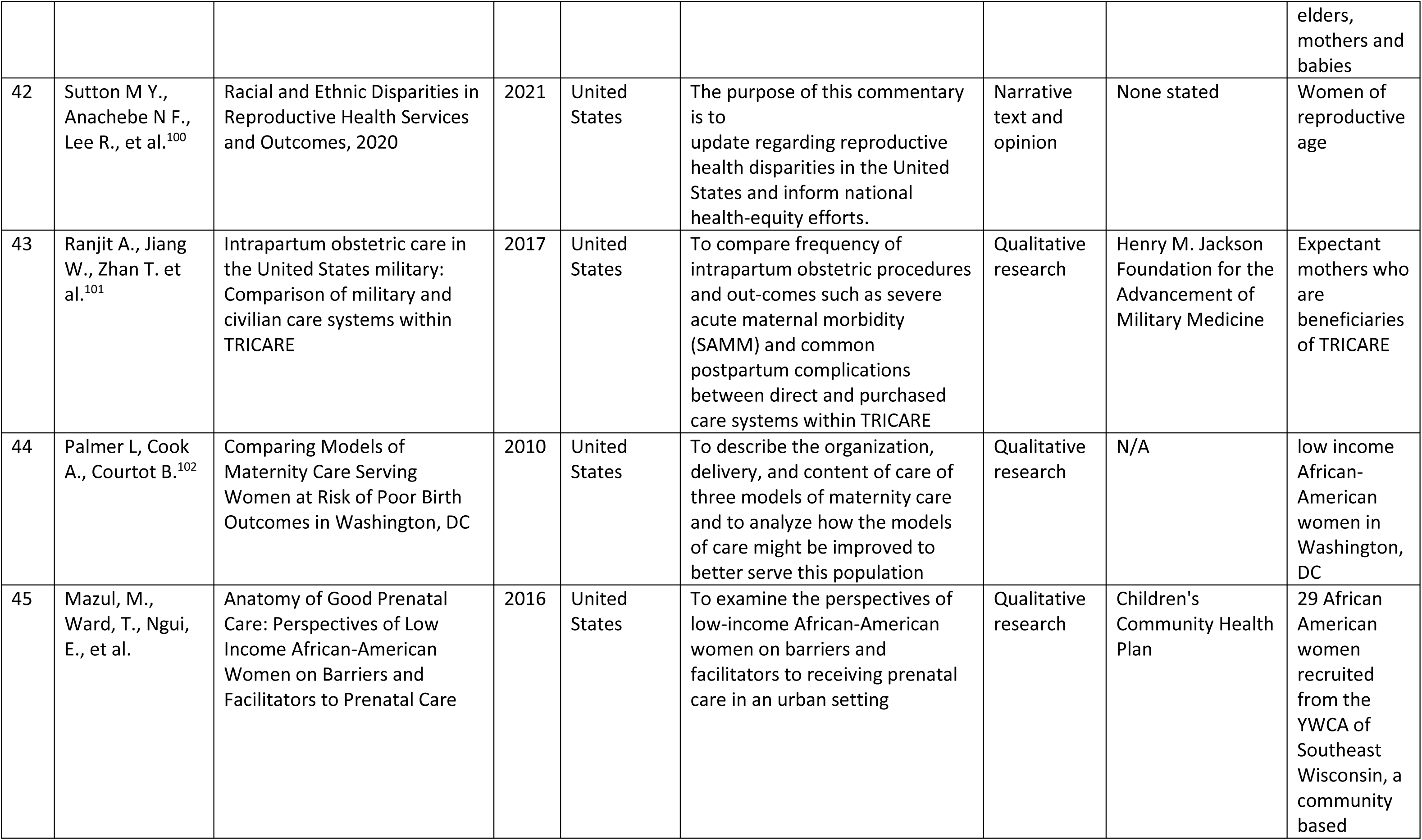
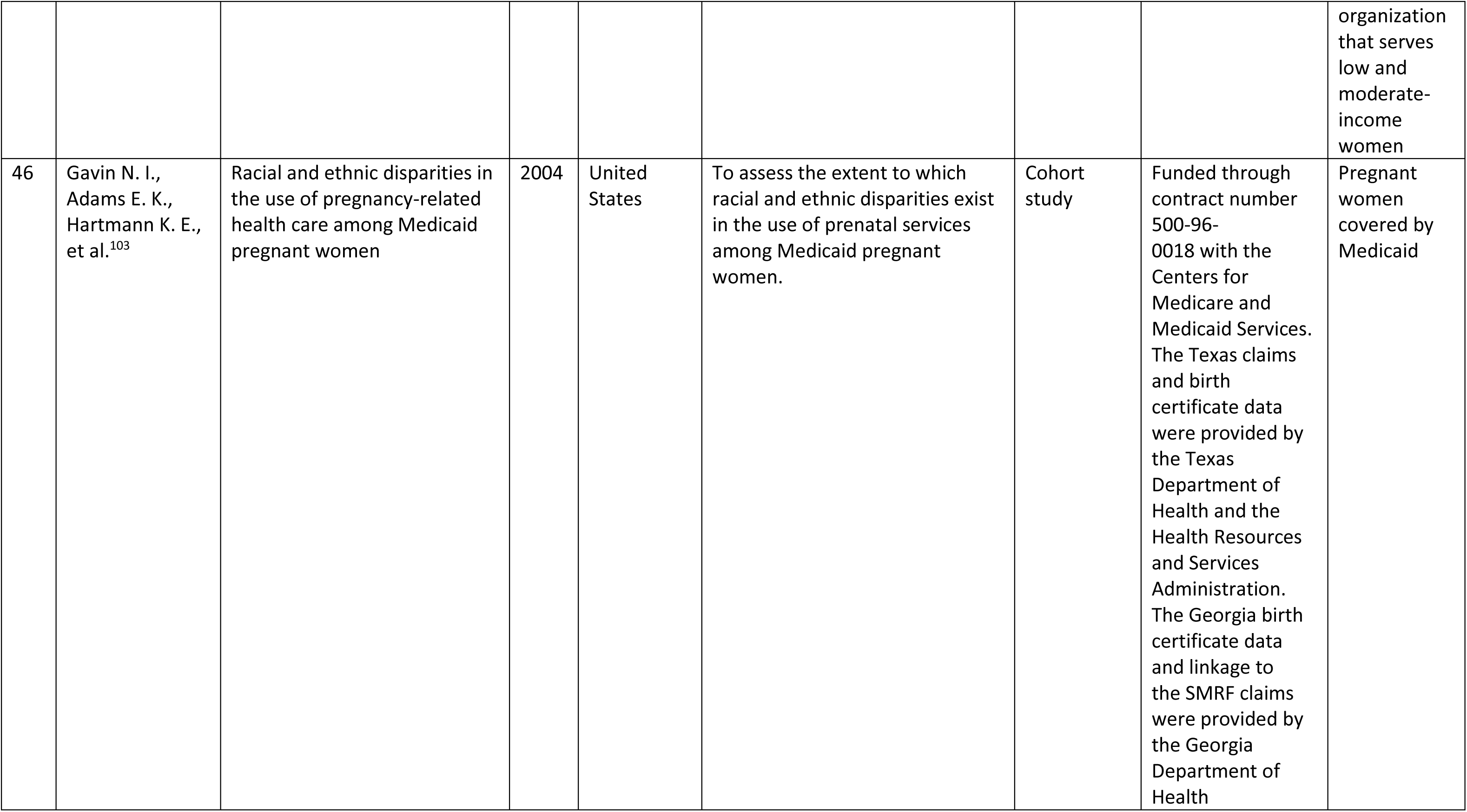
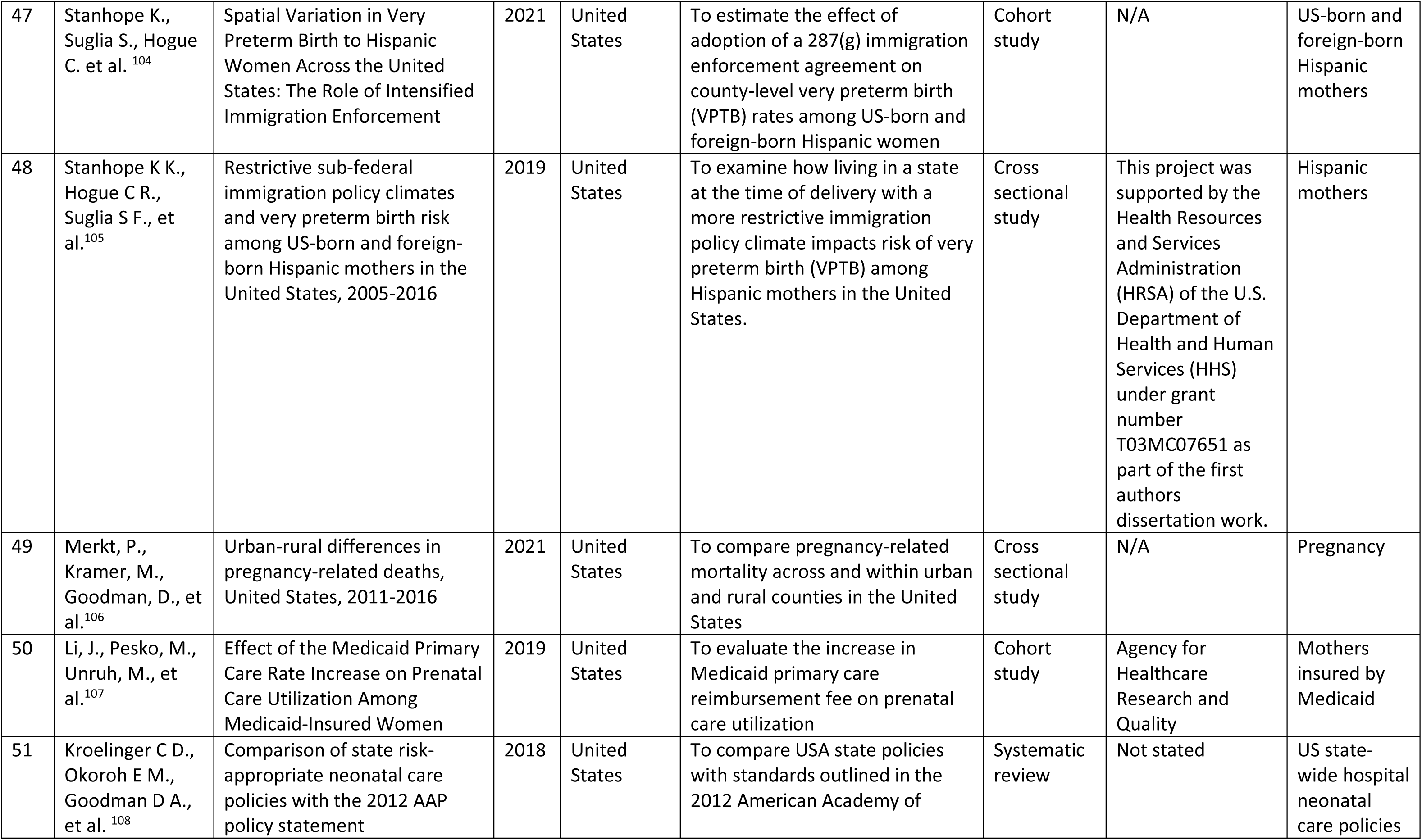
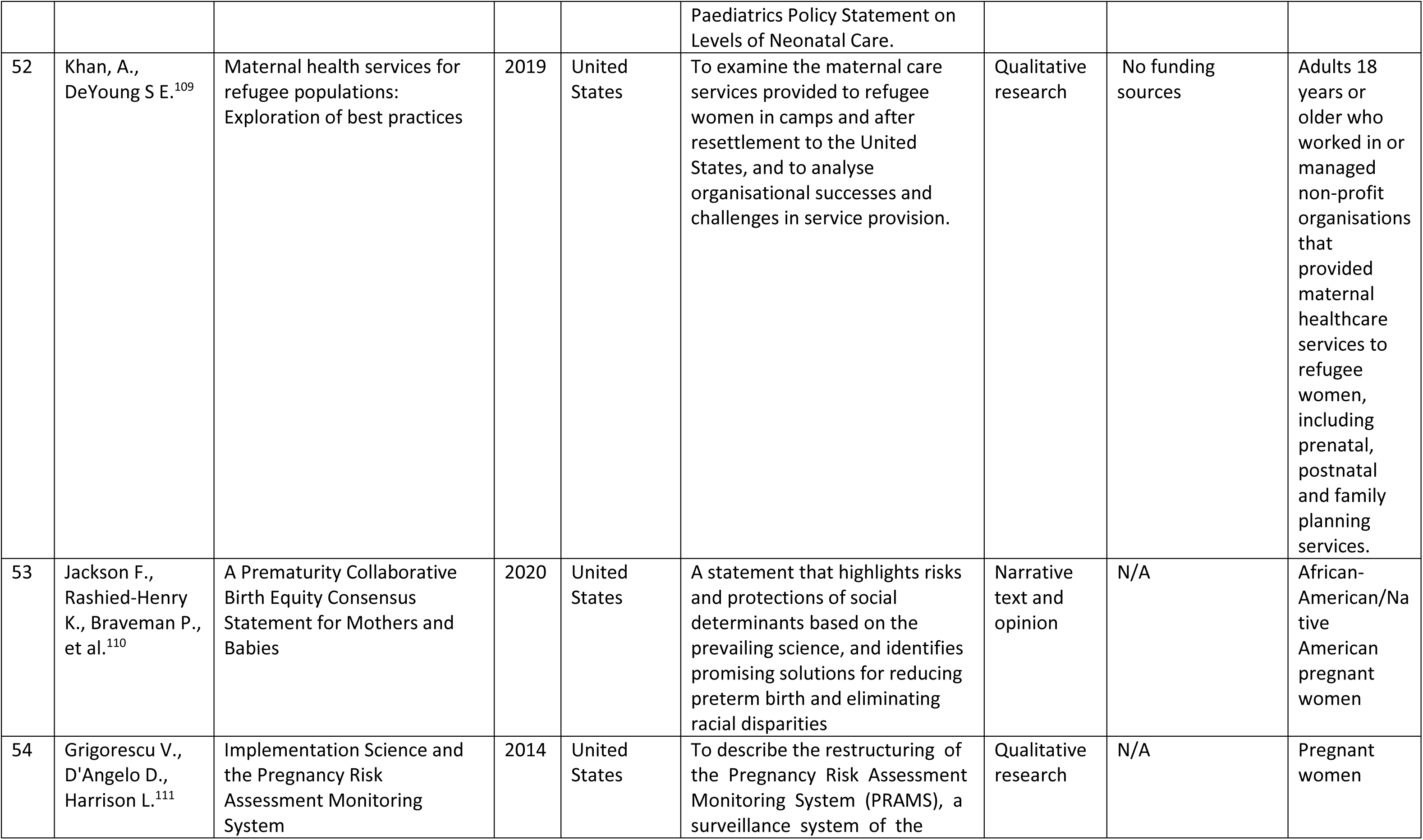
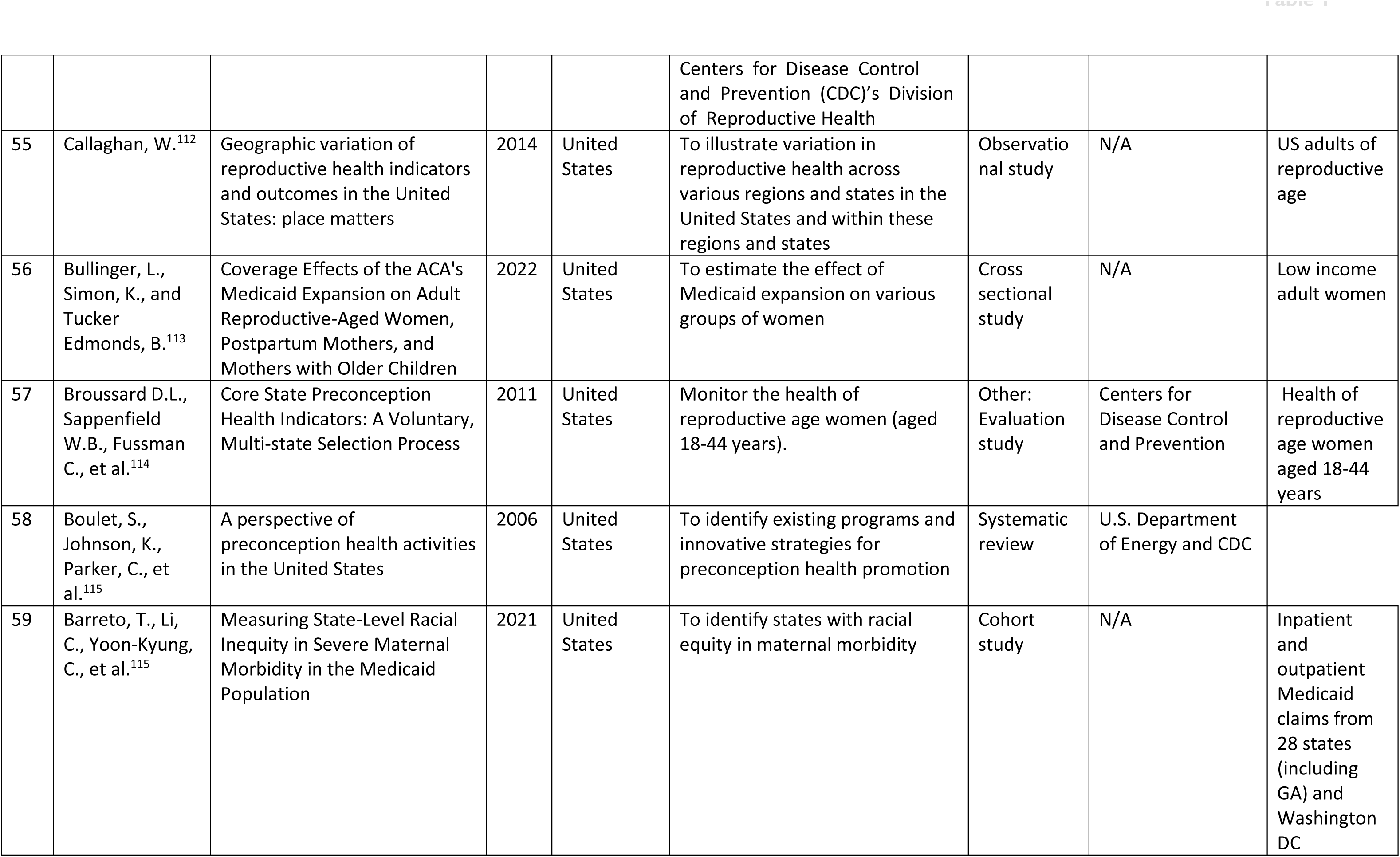
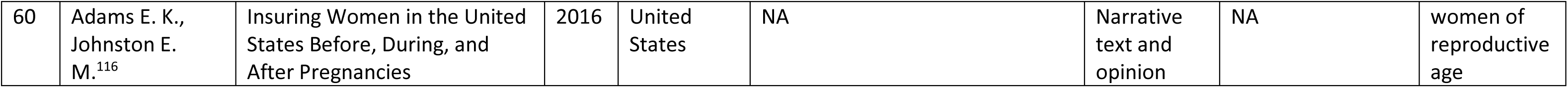
Table of characteristics of included literature.

We carried out a textual synthesis of the data with the intention to review literature that relates synoptically to maternal health service; highlighting similarities and differences between the two countries. The literature outlined the public health systems at three main levels: (1) primary level – action taken at national or country level; (2) secondary level – action taken at policy or legislative level; and (3) tertiary levels – action was taken at regional or a specific city or locality-based e.g., programs.

#### Primary level – national or country level

Maternal care in Scotland is currently provided through the National Health Service Scotland (NHS Scotland).^12^ Scotland is divided into 14 NHS regional Health Boards, each responsible for planning and delivering healthcare services within a specific geographical area.^18^ Specialized maternity services may sometimes be provided at a national or regional level instead of individual Health Boards. Generally, maternal care in the UK includes routine antenatal and postnatal care, midwifery-led care for low-risk pregnancies, and consultant care (obstetrics or specialists care) for higher-risk pregnancies or medical complications.^19^ Scottish professionals in maternity and neonatal care adhere to established clinical and professional protocols that outline the standards for safe and efficient services, developed by organizations like the British Association of Perinatal Medicine (BAPM), National Institute for Clinical Excellence (NICE), and the Scottish Intercollegiate Guidelines Network (SIGN). These guidelines are often protocolized and embedded into practice to support equitable care.

Similarly, the US also strongly adopts evidence-based guidelines and recommendations from the American College of Obstetricians and Gynecologists (ACOG) and Society for Maternal-Fetal Medicine (SMFM) to guide their delivery of maternal and infant care. However, implementation and protocolization of guidelines varies in the US due to its healthcare system, which involves a mix of public and private healthcare providers, insurance systems, and government subsidies.^20^ Healthcare in the US is the responsibility of individual states, and while there are national policies and legislations, it should be noted that there can be significant variations in healthcare planning and policy at the state level. Other healthcare coverage provided by the government includes Medicaid and Medicare which serves different populations. Medicaid serves those whose income and/or resources fall below a designated level while Medicare provides coverage for the elderly.

One of the most heavily cited legislation supporting the expansion of healthcare is the Patient Protection and Affordable Care Act (ACA) that was enacted in 2010. The primary goal of the ACA was to expand access to health and improving the quality and affordability of healthcare.^21^ The historical healthcare reform law mandates that every individual have health insurance starting in 2014. The expansion of Medicaid eligibility under the law has successfully slowed the rise in maternal mortality rates among Black pregnant and birthing individuals in states where it has been implemented. Additionally, the Affordable Care Act (ACA) required the coverage of preventive services, such as contraception, and prohibited discrimination based on pre-existing conditions, including pregnancy. Despite these efforts to expand on health coverage, 26 million people or 8% of the population remains uninsured and rely on safety net programs and charity care.^23^

Furthermore, coverage of pregnancy-related healthcare varies in the state of Georgia by stage of pregnancy.^24^ Medicaid offers access to physicians’ visits, prescription medicines, and in- and out-patient hospital services for pregnant women with an income of up to 220% above the poverty line. Labor and delivery costs are also covered through Medicaid, and coverage lasts up to six months after giving birth. Between 2012-2014, Medicaid was utilized by 12.6% of mothers a month prior to pregnancy while 36.9% of Georgian mothers were uninsured during this time period.^25^

Maternal care models in Scotland and Georgia also vary, with Scotland favoring a midwife-led approach and Georgia predominantly relying on physician-led care.^19,26^ The Best Start report outlined Scotland’s approach to maternity and neonatal care, emphasizing midwife-led models following national guidelines that encourage women without significant health issues to utilize community-based midwife-led care services.^12^ The comparison between midwife-led maternal care in Scotland and physician-led maternal services in Georgia, underscores some of the differences in healthcare models, midwifery roles, and the policies that support the extended scope of practice, in the UK and the US. These variations have been shaped by the historical, cultural, and policy contexts of each region.

#### Secondary level – policy or legislative level

A proportionate number of the included literature were policy and legislative-driven. A summary of the policy and empirical evidence from the textual synthesis can be found in Appendix 2. These policies and relevant frameworks were mapped in chronological order on a timeline for both states as illustrated in Figure 2 and 3.

**Figure 2.**
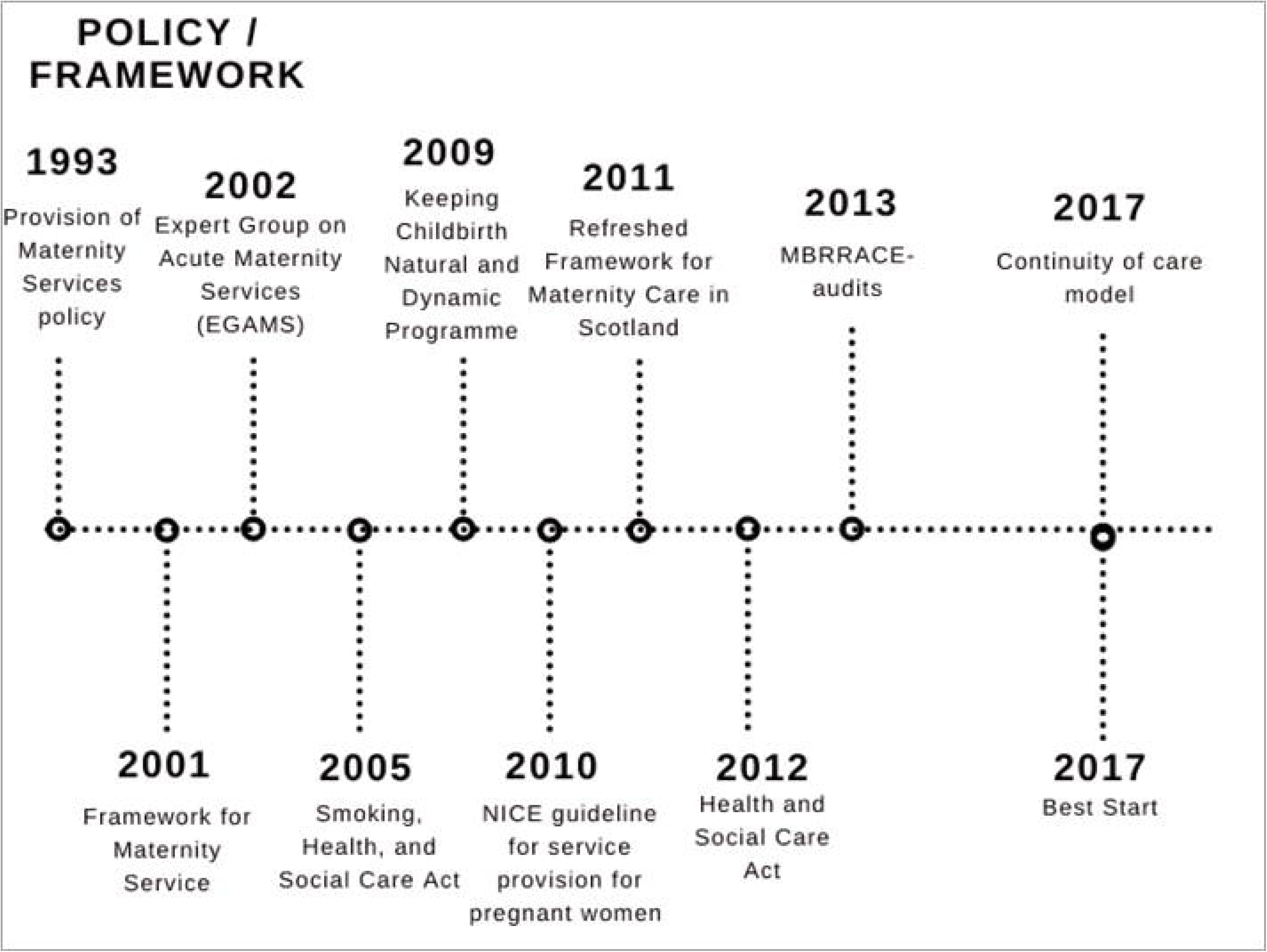
Timeline of maternal policies and legislations in Scotland, UK.

**Figure 3.**
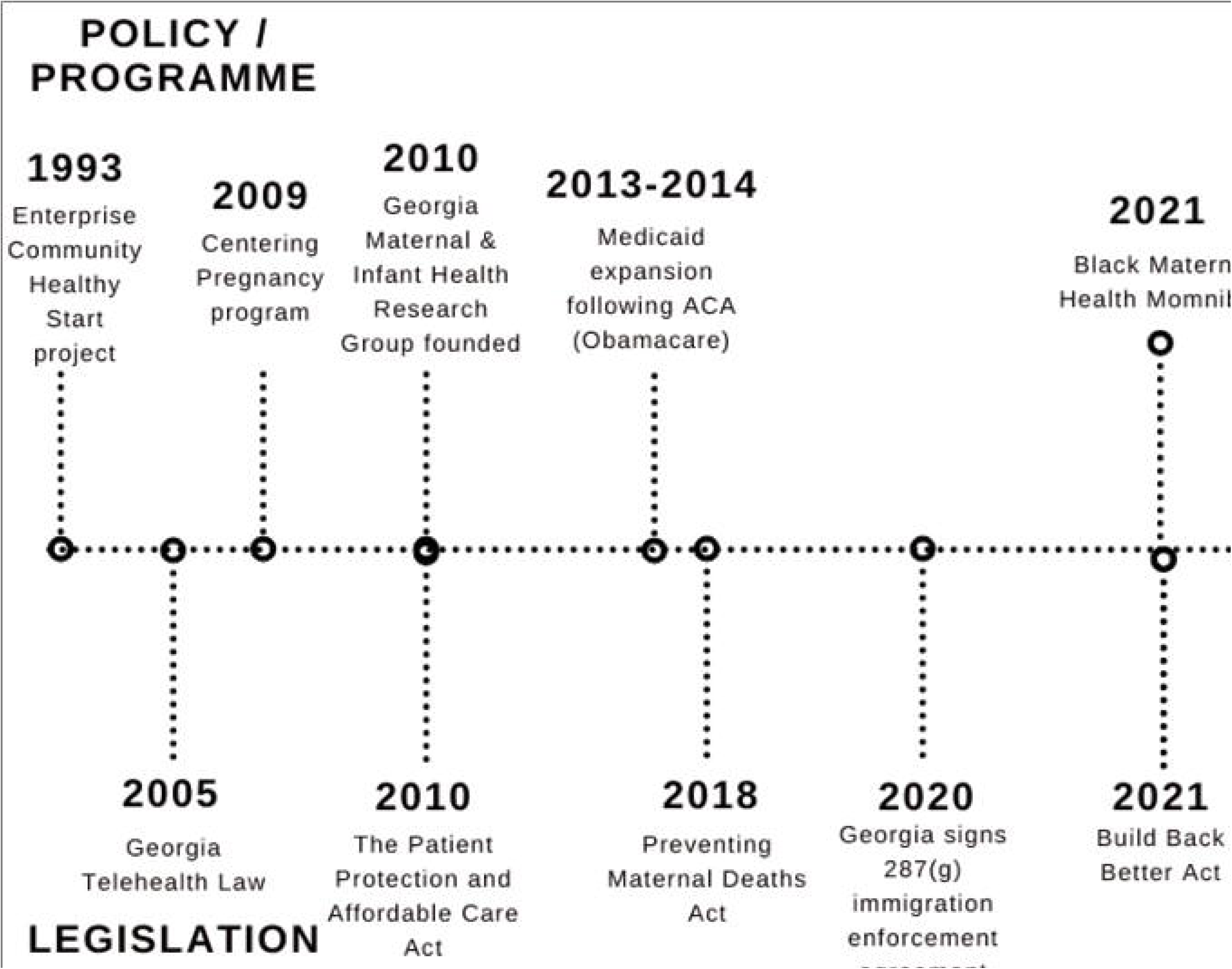
Timeline of maternal policies and legislations in Georgia, USA.

#### Scotland

Several key policy frameworks have played a significant role in shaping the strategic landscape for providing maternity and neonatal services in Scotland. The National Framework for Maternity Services in Scotland^27^ was derived to provide a structured approach to the planning and delivery of high-standard maternity services. Despite the policy’s intentions, historically, there have been translational challenges in practice due to the lack of a clear implementation structure. Expert Group on Acute Maternity Services (EGAMS) was established and published a reference report in 2002 recommending the centralization of maternity services in Scotland and the establishment of interdisciplinary teams to enhance safety and quality of maternity care.^27^ EGAMS also recognized the importance of improving maternity services in rural areas due to the challenges they face, playing a significant role in shaping the direction of maternity care in Scotland. Although the general framework and guidelines for providing maternity care are set at a national level, there is no specific approach or model to service delivery, allowing individual NHS Boards to tailor their services to suit the needs of their locality.

The Refreshed Framework for Maternity Care in Scotland^28^ was further developed to improve maternal and infant health and diminish disparities in health outcomes. It also established guiding principles and service benchmarks for maternity care throughout Scotland. The Quality Framework for Neonatal Care in Scotland^29^ provided guidance on providing high quality, evidence-based, safe, effective and person-focused neonatal care.

Continuity of care had been a pivotal policy affecting maternity care in England since 1993 with the publication of Changing Childbirth and an emphasis on Choice, Continuity and Control, person-centered care in the National Service Framework Maternity Standard and Maternity Matters.^30,31^ Therefore, in 2017, Scotland introduced the ’Best Start’ policy, which identified the future vision of real continuity of care throughout the entire maternity process, with a particular emphasis on supporting vulnerable families as a fundamental aspect in advancing maternity services across Scotland within a five-year timeframe. This current policy emphasizes the provision of consistent care from a dedicated healthcare professional or a team throughout pregnancy, childbirth, and the postnatal period. It promotes person-centered, personalized care and fosters trust between patients and healthcare providers, which has been shown to enhance their overall experience and improve maternal and infant outcomes. NHS Scotland released the Health Improvement, Efficiency, Access to Services, and Treatment Appropriate (HEAT) initiative aimed to measure and improve the timeliness of access to antenatal care for pregnant women in Scotland.^32^ Performance data against current local standards showed that Scotland met the target as the lowest performance even in areas with the highest levels of deprivation (measured by the Scottish Index of Multiple Deprivation^33^ was 88.35% (i.e., 88% of expectant mothers had scheduled antenatal care appointments by the 12th week of pregnancy for the year ending March 2021).

A broad range of ongoing audit, legislative, and improvement efforts have been undertaken with the aim to improve clinical standards and outcomes throughout Scotland. The Maternal and Children’s Quality Improvement Collaborative (MCQIC), initiated in 2013 as part of the Scottish Patient Safety Programme, is one such initiative.^34^ Established in 2011, the Stillbirth Group aims to decrease stillbirth rates in Scotland by increasing awareness of risk factors, supporting research, and advocating for bereavement support and resources.^35^ Another major audit is the Mothers and Babies: Reducing Risk through Audits and Confidential Enquiries in the UK (MBRRACE), established in 2013, which examines maternal deaths, stillbirths, and infant deaths to facilitate continuous quality improvement efforts.^36^ Additionally, the Each Baby Counts program by the Royal College of Obstetricians and Gynaecologists (RCOG) aims to minimize the occurrence of preventable adverse incidents during term labor.^37^ In addition, there are several other public health initiatives that progressed in Scotland to reduce health disparities among diverse populations. These include the WHO self-assessment audit tool to measure the quality of health promotion activity of maternity services in Scotland.^35^ The Smoking, Health and Social Care (Scotland) Act Scottish Government^38^ imposed legislative measures to reduce smoking rates, exposure to second-hand smoke, and smoking cessation support as it recognizes the adverse effects of smoking on maternal and infant health. This legislation resulted in a wide public health impact.^39^

#### Georgia

Several key policies have impacted maternal health care in the state of Georgia, USA. The expansion of Medicaid eligibility for pregnant women has been a critical policy in improving access to maternal healthcare. This policy aims to provide coverage for low-income pregnant women, ensuring access to prenatal care.

Like Scotland, Georgia has implemented policies promoting continuity of care with the same healthcare provider with the aim to establish trust and rapport between healthcare professionals and expectant mothers. The Certified Nurse-Midwife (CNM) Involvement in Care played a vital role in maternal care, offering a more holistic and lower medical intervention approaches to childbirth.

Other programs and initiatives also had positive impacts on maternal and child health in Georgia. Centering Pregnancy: A Model for Group Prenatal Care is a group-based prenatal care model that offers support, education, and a community for mothers-to-be.^40^ Group prenatal care consisted of sessions with a nurse and midwife where basic prenatal physical assessments and issues such as nutrition, common discomforts, labor and delivery, infant care, and postpartum were addressed. Significant collaborations have been established between clinical care, public health, and policy entities at Grady. Examples include initiatives like the Grady Healthy Baby Initiative, aimed at addressing underlying factors contributing to adverse maternal and fetal outcomes, as well as disparities among vulnerable populations, both locally and statewide.^41^

Georgia is also involved in perinatal quality collaboratives aimed at improving the quality of care for mothers and infants. These collaboratives focus on evidence-based practices and guidelines to address maternal and infant outcomes. The Perinatal Case Management programs was introduced in April 2014 to identify and address the complex needs of high-risk pregnant women by providing comprehensive support to reduce the risk of adverse outcomes such as preterm births.^42^ The CDC initiated the Pregnancy Mortality Surveillance System (PMSS) in 1986 to conduct national surveillance of all pregnancy-related deaths. At the state or local level, the Maternal Mortality Review Committees (MMRCs) would convene to investigate and review maternal deaths. These committees were set up to investigate the causes and risk factors for pregnancy-related deaths in the US and develop strategies to prevent future mortalities.

Georgia was one of the first states in the US to adopt telemedicine through the implementation of the Georgia Telehealth Law in 2005. This initiative aimed to establish clear definitions and a legal structure for providing remote medical services to overcome geographical challenges posed by rurality.^43^

#### Tertiary level – regional or specific city or locality-based level

Different models of care delivery have been implemented in Scotland and Georgia including traditional physician care, group-based prenatal care, and midwifery-led care. The overarching goal is to provide person-centered care and higher satisfaction among expectant mothers.^44^ All pregnant women will see a range of health professionals depending on their care needs. While the majority of births occur in hospitals, the choices available for birthplace can vary depending on the locality and are often influenced by the individual preferences of the woman. In Scotland, approximately 2.6% of births occur in community settings, and antenatal care can be provided in both community and hospital settings. In the United States, nearly all births (98%) take place in hospitals,^45^ with approximately 91% attended by physicians and 8.7% attended by midwives, a statistic that is unique to the US.^46^ In contrast, many other high-income countries rely more heavily on midwifery care and have fewer hospital births.^47^

Every maternity unit in Scotland has obtained accreditation from the UNICEF Baby Friendly Initiative (BFI).^48^ Scotland’s four largest neonatal units are close to achieving full implementation of the neonatal BFI standards, while other units are working towards this goal. Additionally, a Scotland-wide donor milk bank was established in 2013 to ensure equitable access to breast milk for the smallest and most vulnerable infants across the country.^49^ In Georgia, the involvement of peer counsellors, particularly in breastfeeding support programs has been beneficial in encouraging and supporting breastfeeding which has documented health benefits for both mothers and infants.

## Discussion

Our findings demonstrate the varied approaches of the Georgia and Scotland health systems and their influence on maternal care and wellbeing. Both systems share a commitment towards providing continuity of care for the expectant mothers in their regions, acknowledging the clinical benefits of promoting deepened person-centered trust. Their differences originate at a national level resulting in macro-level barriers and facilitators to health (e.g., availability of infrastructure, facilities, and medical staff).^50^ Scotland has a heavily guideline-driven approach to its medical practice due to its centralized health system framework, as opposed to the largely privatized US healthcare market, which subsequently allows additional room for personalized care. Furthermore, the US healthcare system is more physician-centered compared to its Scottish counterpart, which prioritizes midwifery-centered care in the absence of high-risk comorbidities or complications. This discrepancy in practitioner emphasis allows the differences to pervade into the regional level, as demonstrated by the variations in delivery setting: births in the non-hospital setting are not uncommon in Scotland, while the vast majority of births in the US occur in physician-led hospitals.

Georgia has one of the more restrictive state policies regarding the licensing of midwives, with the criminalization of midwifery practice without a nursing credential. As of 2018, there are only 550 Certified Nurse Midwives (CNMs) in the state of Georgia^51^ and they face challenges such as identifying physician collaborators and obtaining hospital privileges, making it extremely challenging to access midwifery care. Additionally, 73% of counties in Georgia lack hospitals that provide maternity care, with 36.7% being classified as ’maternity care deserts’.^52^ In response to this maternal health crisis, Georgia has implemented several initiatives, including a Maternal Mortality Review Committee (MMRC), and a Perinatal Quality Collaborative (GaPQC) involving key stakeholders.

The demographic and geographic variations between Georgia and Scotland may also explain their varied policies. Georgia’s estimated 2022 population is 10.9 million. While most of the inhabitants are White (59%), 33% are Black/African American, 10% are Hispanic, and 4.8% are Asian. A significant portion of the state’s population, 39.7%, live in rural parts of the state with higher poverty rate compared to urban areas of the state (19% *vs.* 13%) based on American Community Survey (ACS) data.^53^ Scotland, by comparison, has a smaller population of 5.4 million, with only 17% of its population living in rural areas of the country.^54^ The racial breakdown of Scotland’s population is more homogenous to that of Georgia, with 96% of its population being White.^55^ Such variations in spatial barriers and socioeconomic status are important when considering the differences in birth outcomes. The larger, more diverse, and more rural population in Georgia can present with more obstetric and neonatal challenges, leading to a higher need for maternal care services.

The different healthcare infrastructures present in Georgia and Scotland may also be due to the conceptual framework underlying their respective public health systems and the populations each system was originally designed to serve. The US public healthcare system, through programs known as Medicare and Medicaid, was established in the 1965 to provide care to the elderly and others who were deemed medically and financially disadvantaged in the setting of rising healthcare costs.^56^ Despite these programs, the US healthcare system continues to be driven by the private sector, with employer-based coverage currently being the main source of health insurance for working families.^57^ As such, social determinants of health including economic stability, timely access to health care providers and the quality of maternal care are known risk factors for maternal outcomes.^4^ A recent survey in Georgia revealed that improving the affordability of maternal health care and access to insurance coverage remain the top priorities to tackle this health crisis.^58^ On the other hand, the UK National Health Service (NHS) was implemented in 1948 to provide preventative and curative healthcare to its entire population after the nation endured large numbers of its population requiring medical attention during World War II. Therefore, with the UK’s emphasis on public healthcare coverage for a broader target population, Scotland’s inhabitants benefit from a more accessible and affordable healthcare system compared to their American counterparts.^59^

This study is the first to comparatively explore health policies between Georgia and Scotland, two regions with vastly different healthcare systems. Furthermore, no studies explore varied policies between the broader US and UK health systems. Our study utilizes primary literature, such as clinical guidelines and legislative documents, and secondary literature, such as news articles and expert opinions. While this allows us to gain a holistic insight through objective information regarding Scotland and Georgia’s policies as well as subjective information regarding their impact on their respective communities, the use of grey literature such as news and expert opinions may introduce some bias. One strength is our multi-layered approach to policy impact, allowing us to clearly demonstrate how healthcare policy has a trickle-down effect on the provision of healthcare services from the national to the local level. However, the study’s focus on maternal policies may overlook other important factors such as individual health behaviors and community-level determinants of health. Furthermore, the study was limited by the lack of literature on rural health policies in Scotland which demonstrates decreased rurality-centered policy in Scotland, representing an area for benchmarking.

The results of this study have potential to shape approaches to maternal care and can be utilized to provide recommendations for future clinical practice in Georgia and Scotland. For example, staffing shortages have long been a contributor to the maternal mortality rate in Georgia.^24^

Scotland was able to address this concern through increased mid-level provider involvement in uncomplicated cases, which increases access to basic prenatal needs across the country and subsequently boosts obstetrician availability for more complex cases (Scottish Government, 2017).^12^ These findings align with current discussions regarding licensure for midwives and maternity care providers, reflecting the timeliness and relevance of this textual synthesis. The evidence suggest that an expansion in midwifery licensure and training, extended scope of practice, and integrated community-based practice is equitable and cost-effective in reducing the gap in maternal health.^47,51,52,60^ While these changes may affect policy at the regional level, broader changes at the legislative or national level would require fundamental changes to their respective socioeconomic and healthcare system.

## Conclusion

This study has identified an important gap in literature addressing the health challenges faced in rural Scotland. This information is necessary to understand the current operations and challenges of antenatal and maternal care in those areas and to provide an important area of comparison to Georgia. Additional research, incorporating population-based data, is needed to fully understand the impact of policy on maternal and child health outcomes in rural Georgia and rural Scotland. It is also important to explore additional indicators of healthcare quality, including patient satisfaction and the availability of healthcare resources.

## Data Availability

All data produced in the present work are contained in the manuscript.

## Ethics statement

No ethical or IRB approval was needed because this is a study of secondary data evidence synthesis.

