## Supplementary Files for "Understanding the impact of antenatal care policies in Georgia (USA) and Scotland (UK): A textual synthesis"

Appendix 1: Search strategy

The following logic grid was proposed to develop the search strategy for this review:

| Population (OR) | Phenomenon of Interest (OR) | Context (OR) |
| --- | --- | --- |
| Maternal health  Maternal mortality  Maternal morbidity  Perinatalcare  Prenatal care  Postpartum care  Antenatal care  Antepartum care  During pregnancy care  Antenatal  Pregnancy  Pregnant  Prenatal  Perinatal  Maternal | Polic* *(to include policy, or policies)*  Legislative  Legislation  Framework  AND | United Kingdom  UK  Scotland  Scottish  Georgia  America  United States  USA  AND |

Appendix 2: Table of findings

| **Authors** | **Year** | **Country in which the study conducted:**  **UK= 3**  **Scotland = 12**  **Global= 5**  **Georgia= 18**  **USA= 21** | **Significant results/findings of textual review** |
| --- | --- | --- | --- |
| Puthussery S. | 2016 | UK | The UK has attempted to implement policies that address health inequities in migrant women. However, the integration of migrant health into broader policies and the fluid rules in some of these policies has to led some ambiguity and confusion for healthcare providers and healthcare provision as a whole. |
| Courtemanche, C., Marton, J., Ukert, B., et al. | 2017 | United States | Georgia is a non-expansion state, which based on the context of this study, indicates lower gains in insurance coverage compared to expansion states |
| MacIver E., Lau, A. | 2019 | Other: Conducted in Scotland but wide scoping review | Limited significance as post birth but key transferable point may be as follows: women need consistent information and advice and value personalised care. Models of care that facilitate these needs are focused on relational continuity and lead to greater satisfaction. |
| McInnes, R., Aitken-Arbuckle, A., Lake, S., et al. | 2020 | Scotland specific | CMC models have shown to be effective, but necessitate effective leadership for sustainable implementation in Scotland. |
| 244 American Health and Care organisations -led by American College of Obstetricians and Gynaecologists (listed first) | 2021 | United States | The Build Back Better Act includes robust investments in programs that take steps towards birth equity by centering the voices, needs, and preferences of Black and Indigenous individuals, who disproportionately experience inequities in maternal health. Inclusion of the Black Maternal Health Momnibus Act focuses on investing in community-based organizations, addressing social determinants of maternal health, expanding the perinatal workforce, addressing the impacts of climate change on maternal and infant health, increasing community engagement in maternal mortality review committees, supporting targeted research at Minority-Serving Institutions, investing in maternal mental health, and improving the provision of culturally congruent care |
| Abraham G. (President of American college of Physicians) | 2021 | United States | The BBBA is supported by the ACP (the largest medical specialty organization and the second-largest physician membership society in the United States), in addressing paid leave/federal paid maternity leave, maternal health, mortality and behavioural Health care expansion. Medicaid expansion also mentioned but in postpartum context. |
| Till SR, Everetts D, Haas DM | 2015 | Other: Globally | - Women who received incentives were no more likely to return for postpartum care.  - Women who received incentives were more likely to attend prenatal visits on a regular basis and obtain adequate prenatal care (i.e. vaccinations, blood pressure readings, breastfeeding counseling). |
| Turienzo C., Bick D., Briley A. et al. | 2020 | UK | The number of live-born infants and fetal losses before 24 weeks' gestation did not statistically differ between the group of expectant mothers with continued midwifery care and the group that did not. |
| Gale N.K., Kenyon S., MacArthur C., et al | 2018 | UK |  |
| Vladutiu, C., Mobley, S. Ji, X., et al. | 2021 | Georgia specific |  |
| Vanderlaan J., Edwards J., Dunlop A. | 2019 | Georgia specific |  |
| Sutton M Y., Anachebe N F., Lee R., et al. | 2021 | United States | Racial-ethnic disparities in reproductive health access, services, and outcomes are prevalent and require heightened awareness and strategies to close these long-standing disparity gaps. Specifically, for preventing maternal deaths, developing and strengthening policies and laws that include a focus on dismantling structural racism and implicit bias are crucial as part of the solution.  Relevant components of the Affordable Care Act (ACA) and the Preventing Maternal Deaths Act could help to further strengthen reproductive health care, close gaps in services and outcomes, and decrease racial-ethnic reproductive health disparities. |
| Rust G., Nembhard W., Nichols M, et al. | 2004 | Georgia specific | - Epidural analgesia was provided for 62.1% of births in urban/metropolitan counties versus 39.2% for rural/non-metropolitan counties. - Our study found significant racial/ethnic differences in the rates of epidural analgesia for the management of pain for those receiving Medicaid. |
| Ranjit A., Jiang W., Zhan T. et al. | 2017 | United States | In stratified analysis by mode of delivery, purchased care had lower odds of common complications for all modes of delivery. |
| Pinto M., Rochat R., Hennik M., et al. | 2016 | Georgia specific | To provide public health officials, policy makers, and programmatic administrators the opportunity to combat Georgia's high maternal and infant mortality rates, core principles such as continuity of care and collaboration with advanced care practitioners should be implemented within the Georgia healthcare system. |
| Palmer L, Cook A., Courtot B. | 2010 | United States | Adopting group prenatal care and increased use of CNMs or NPs may increase efficiency and improve provision of nonclinical care. |
| Mosley E A., Pratt, M., Besera G., et al. | 2021 | Georgia specific | Community-based, culturally tailored pregnancy support programs like Embrace can meet the complex needs of refugee women. Additionally, community-engaged, cross-sector research approaches could ensure the inclusion of both community and clinical perspectives in research design, implementation, and dissemination. |
| Mobley S., Dixson Thomas S., Sutherland D., et al. | 2014 | Georgia specific | There was a progression in the MHL of rural, low-income mothers through case management; namely women were 57 times more likely to score as adequate on the final postpartum LSP/HCL and 3 times more likely to score as adequate on the LSP/SCL than the prenatal assessments. |
| Meyer, E., Hennink, M., Rochat, R., et al. | 2016 | Georgia specific | Strong patient-provider communication showed to be essential in receiving quality care. Participants consistently expressed delays and anxieties related to the Medicaid process. Further, participants who received Medicaid benefits described feeling less worthy of aspects of the healthcare system. Several key informants noted their clients lacked reliable sources of information on pregnancy and the need for adequate and timely prenatal care. Feelings of shame and stigma, particularly amongst adolescents have shown to delay care, as described by key informants. Women in shortage areas reported facing more difficulties locating providers with their preferred criteria. |
| Mazul, M., Ward, T., Ngui, E., et al. | 2016 | United States | 1. Structural barriers were described as the most prominent barriers. These barriers include difficulty acquiring and maintaining Medicaid, difficulty finding providers that would accept their insurance, transportation to prenatal care appointments, inflexible clinic hours, and long wait times. 2. Psychosocial barriers included poverty, legal issues, homelessness, family relationships, lack of social support, and experiences of discrimination. 3. Unintended pregnancy, perceptions of poor quality of care, and perceptions of discrimination also prevented women from attending their prenatal care visits. |
| Grant J H., Handwerk K., Baker k., et al. | 2018 | Georgia specific | CenteringPregnancy administered via public-private partnership may improve access to prenatal care and perinatal outcomes for medically underserved women in low-resource settings e.g: Since the inception of CenteringPregnancy, Georgia’s Southwest Public Health District (SWHD) has consistently initiated prenatal care within 2 weeks of appointment requests - external funding enabled removal of two known barriers of early prenatal care access in low-income women insurance status and transportation. Initiation of prenatal care regardless of Medicaid enrolment status, three out of four pregnant patients at the Dougherty County Health Department (part of SWHD) initiated prenatal care in the first trimester. |
| Gavin N I., Benedict M B., Adams E K. | 2006 | Georgia specific | The results indicate that there could be chances to enhance the availability of prenatal care for Medicaid beneficiaries who qualify for blind and disabled categories. (NB - they do not suggest how that may be accomplished) |
| Gavin N. I., Adams E. K., Hartmann K. E., et al. | 2004 | United States | Unexplained racial and ethnic disparities persist in the initiation and use of both routine and specialised prenatal care services among women in Georgia who are fairly homogeneous with respect to socioeconomic status and who are provided equal financial access to health care services. This suggests disparities arise from both patient- and provider-related factors.  Local community outreach programs familiar with the cultural expectations of minority groups and provider education programs to raise the cultural awareness and sensitivities of providers may be needed to eliminate racial and ethnic disparities |
| Daymude A E C., Daymude J J., Rochat., R. | 2022 | Georgia specific | Rural LDU closure in Georgia has a disproportionate impact on Black and low-income women and may be prevented through funding maternity healthcare, financing LDUs, and addressing provider shortages |
| Bruce F C; Berg C J.; Joski P J., et al. | 2012 | Georgia specific | 1. Comprehensive health insurance coverage may lessen the unfavourable impact of socio-economic disadvantage on the risk of maternal morbidity. 2. Compared with all other race/ethnic groups, a greater proportion of pregnancies of Black women were in the lowest neighbourhood SES group and were associated with at least one complication.  3. Even for White women, the effect of SES is small, suggesting that the degree to which SES influences the risk of morbidity may be ameliorated by comprehensive health insurance coverage NB The authors acknowledge that residual confounding may have accounted for the associations observed and suggest caution in the interpretation of the findings |
| Barkin J. L., Bloch J.R., Smith K.E. R., et al. | 2021 | Georgia specific | Home visiting could confer substantial benefit to women with high-risk obstetric conditions who often require greater support than the general pregnant and postpartum population. However, some may feel uneasy with the idea of allowing visitors in their house. For home visiting programs to effectively reach this population, women need more education about how home visiting programmes function and what purpose they serve, in addition to assurances related to safety, privacy, and boundaries. Additionally, health care providers could suggest alternative resources, such as support groups or other local social programs, if individuals are reluctant to participate in home visiting. Clear communication is needed regarding the functions of home visiting programs to dispel apprehension |
| Armstrong-Mensah, E., Dada, D., Bowers, A., et al. | 2021 | Georgia specific | Between 1994 and 2015, there has been a significant amount of hospital closures throughout the state of Georgia. In 2019, 93/109 rural counties lacked labor a labor and delivery unit with 75 lacked OB/GYNs. Poor communication with providers has been linked to delayed and discontinued care, with is also significant in women with low SES. Georgia's decision to not expand Medicaid has affected access to prenatal care. 21% of African American women in Georgia experience some form of racism before, during, and/or after birth. |
| Zertuche, A., Spelke, B., Julian, Z., et al. | 2016 | Georgia specific | When adjusted for cost inflation, Medicaid reimbursement for obstetric care declined by 37% from 2001 to 2011. There are over 30 counties throughout the state of Georgia without OB services and many more with a deficit in providers. Provider narratives show malpractice costs, underfunded patients, and an overwhelming quantity of patients and deliveries contribute to difficulties in recruiting providers. |
| Young, D., Shield N., Holmes A., et al. | 1997 | Scotland specific | Women in the midwife-managed group were significantly more satisfied with the accessibility of antenatal care compared to those who received shared care. |
| Stanhope K., Suglia S., Hogue C. et al. | 2021 | United States | - There was no global effect of 287(g) adoption on county VPTB rates. - In counties in the SE region of the US, 287(g) adoption was associated with increased county VPTB rates among foreign-born Hispanic women. |
| Stanhope K K., Hogue C R., Suglia S F., et al. | 2019 | United States | Living in a state with more restrictive immigration policy climate was associated with a slight increase in the odds of very preterm birth (after controlling for individual, county, and state level)  State level lagged, Immigration Climate Index (ICI) predicted additional risk of VPTB among Hispanic women. |
| Pitchforth E., Watson V., Tucker J., et al | 2007 | Scotland specific | In contrast to service redesign offering local midwife-managed intrapartum care, most rural women in our study expressed a preference to give birth in hospital and have consultant-led care because they felt safer. |
| Pitchforth E., van Teijlingen E., Watson V., et al. | 2009 | Scotland specific | The findings have direct relevance for maternity services through an understanding of how women may prioritise between different models of care (also for what is understood by the concept of “choice” in the health service as a whole). Provision of different models of maternity services may not be sufficient to convince women they have “choice”. Fundamental questions remain about the meaning of “choice” within current policy developments. The paper calls for a more critical approach to the use of choice as a service development and analytical concept. Further work is needed to understand the role of choice for women and how this can most usefully inform ongoing service redesign. |
| Merkt, P., Kramer, M., Goodman, D., et al. | 2021 | United States | Non-Hispanic Black women had higher mortality rates than non-Hispanic White women in all urban-rural categories, with the disparity ratios being 3-4 times higher. In small metro, micropolitan, and noncore counties, mortality ratios of non-Hispanic Alaska Native and American Indian women were 2 to 3 times higher than that of non-Hispanic White women. Non-Hispanic Asian or Pacific Islander women had a 1.3 times higher ratio in large metro counties compared to non-Hispanic White women, but this disparity is not statistically significant in other categories. |
| McInnes, R., Hollins Martin, C., and MacArthur, J. | 2018 | Scotland specific | Internationally, there are inconsistent findings on the impact of MCC models on the work-life balance of midwives. Evidence suggests involvement and negotiation between midwives and management is essential for successful implementation of the MCC model. |
| McGuire, M., Dagge-Bell, F., Purton, P., et al. | 2004 | Scotland specific | There are 22 consultant-led maternity units in Scotland that perform 180-6000 births per year and 18 maternity units that perform less than 10 to 180 deliveries per year. The Framework for Maternity Services in Scotland was launched in 2001 to promote the normality of pregnancy and women and family centered, community-based, and midwife-managed care. This article notes that increased specialization and restrictions on working time make previous medical staffing untenable during intrapartum and neonatal care. Particularly, difficulties in recruitment and retention and increased demands of clinical governance make it difficult to provide properly trained workforce for intrapartum care. The Expert Group on Actute Maternity Services (EGAMS) published a report that highlights these difficulties, leading to reviews of maternity services throughout Scotland. |
| MacLachlan A., Crawford K., Shinwell S. | 2021 | Scotland specific | A change in maternity policy occurred in 2016, enforcing all women to attend sonography clinics at 'maternity hubs’ (maternity hospitals offering central services for antenatal clinics, scan clinics and maternity wards) rather than more local maternity hospitals or community clinics. This provided THRIVE researchers with a wider pool of women at one location and a scenario where all women had a viable pregnancy. This ultimately resulted in an increase in the number of referrals generated per month, particularly from maternity hubs. |
| Li, J., Pesko, M., Unruh, M., et al. | 2019 | United States | Georgia is considered a large fee bump state so the benefits of the fee bump likely translate to Georgia. In addition, this study controls for Medicaid expansion (Georgia is a non-expansion state) and found it was not a confounding variable. |
| Lanier, P., Kennedy, S., Snyder, A., et al. | 2022 | Other: 6 Southern US States, including Georgia | Georgia is a non-expansion state where 46.1% of births are covered by Medicaid. State law as of August 2021 requires screening at first prenatal visit and third trimester visit. Screening is also required at delivery if there is no prior screening (as per Georgia code 31-17-4.2). Between 2018-2019 63.6% of Medicaid enrollees residing in urban areas received screening and 71% of enrollees residing in rural areas received screening. |
| Kroelinger C D., Okoroh E M., Goodman D A., et al. | 2018 | United States | Neonatal focus so significance limited - state policies vary in consistency, which is a potentially significant barrier to monitoring, regulation, uniform care provision and measurement, and reporting of national-level measures on risk-appropriate neonatal care. |
| Kramer, M., Waller, L., Dunlop, A., et al. | 2012 | Georgia specific | Following the closure of public housing settlements, women residing in these areas were forced to make housing transitions. Compared to women who were not forced to make this move, women who were forced to make this move had an elevated risk of preterm low birth weight. |
| Khan, A., DeYoung S E. | 2019 | United States | This study provides insight into what services are provided to women in refugee camps and upon resettlement in the United States. Individualised education and connection to medical services is prominent, with an emphasis on working with community liaisons and/or multilingual staff to promote communication and trust. Challenges to service delivery include: the need for increased personnel, physical resources, funding, and political support.  Addressing these barriers would allow non-profit organisations to continue to provide culturally and linguistically appropriate maternal health and social services to refugee women. |
| Jincharadze N., Kazakhashvili N., Sakvarelidze I. | 2018 | Georgia specific |  |
| Jamieson, D., Haddad, L. | 2018 | Georgia specific | There are currently several maternal health initiatives in the state of Georgia, but most are centered around Atlanta and Grady Memorial Hospital. Initiatives include expanding access to contraceptives and family planning services, the Grady Healthy Baby initiative (relieves socioeconomic barriers to antenatal care), and group pregnancy counseling. |
| Jackson F., Rashied-Henry K., Braveman P., et al. | 2020 | United States | N/A |
| Hundley, V., Rennie, A., Fitzmaurice, A., et al. | 2000 | Scotland specific | New policies in Scotland aim to increase autonomy of women over their pregnancies, but there is still a gap in implementation of these policies. |
| Hundley V., Rennie A., Fitzmaurice, A., et al. | 2000 | Scotland specific | The paper suggests that based on the survey results, considerable efforts have been made to improve information and choice for women however it also observes that further work is needed if women are to be offered informed choice in the provision of their maternity care e.g.: Improved access to information is essential if women are to have real choice Written information may facilitate choice (the majority of women reported receiving the recommended pregnancy and childbirth book). The fact that nearly a third of women did not know that they could have all their care from midwives and almost half did not know what a DOMINO delivery was, clearly indicates that these methods of providing information do not always convey the available options for care |
| Hundley, V., Penney, G., Fitzmaurice, A., et al. | 2001 | Scotland specific | Scotland has implemented several policies to improve antenatal care, including distribution of a standard pregnancy book, creating models for adequate amount of antenatal visits, providing birth plan autonomy for mothers, establishing reunion groups, providing an identifiable maternity care co-ordinator, and providing continuity of care. Although these policies have been implemented, there are discrepancies in whether mothers receive these interventions and whether providers are aware of these discrepancies. |
| Harron, K., Verfuerden, M., Ibiebele, I., et al. | 2020 | Other: Scotland, England, New South Wales, Ontario, and Sweden | Out of all 5 countries, Scotland has the highest teen pregnancy rate and highest number of infant deaths among teen mothers |
| Harron, K., Verfuerden, M., Ibiebele, I., et al. | 2020 | Other: Scotland, England, New South Wales, Ontario, and Sweden | Out of all 5 countries, Scotland has the highest teen pregnancy rate and highest number of infant deaths among teen mothers.  (note: Infants born to mothers aged <25years represent a large proportion of births in England and Scotland). Although policies and primary pre-ventive strategies have been suggested to reduce and postpone teenage pregnancies, it is not implied that it is current practice in Scotland. England has introduced a multifaceted policy intervention involving health and education agencies has contributed to a decline in teenage and appears to have attenuated the steep deprivation gradient. |
| Grigorescu V., D'Angelo D., Harrison L. | 2014 | United States | According to data pulled from the PRAMS sites with a sufficient response rate, 90.9% of sites in Georgia do not perform a pre-pregnancy blood pressure check for preconception health. |
| Frank J., Bromley C., Doi L., et al. | 2015 | Scotland specific | 1. The extent of provision, throughout the UK, of universally accessible (free at point-of-care), prenatal and perinatal services is more favourable than many developed countries (Roberts, 2012). Current UK-wide services through the National Health Service (NHS) ensure that virtually all mothers and children have very good chances of receiving, at no direct cost, high-quality prenatal/perinatal care by international standards, according to their needs and preferences, regardless of their gender, place of residence, ethnicity or socioeconomic position (SEP) (Krieger et al., 1997), thus enhancing equitable health outcomes (Marmot et al., 2008; Schoen et al., 2010). For example, there is little variation in the levels of expected mortality among very preterm babies of different socioeconomic backgrounds, receiving similar neonatal care (Smith et al., 2009). Nevertheless, infants born into social disadvantage in the UK continue to experience adverse birth and infant outcomes, including low birth weight, premature birth, stillbirth, and infant mortality |
| Cheyne, H., Abhyanker, P., and McCourt, C. | 2013 | Scotland specific | The Scottish Government's support of normal birth through midwife-led care and the reduction routine intrapartum intervention showed to be successful |
| Callaghan, W. | 2014 | United States | In 2012, Georgia had the 13th highest teen birth rate. Infant mortality rates in Georgia are consistently higher for Black infants compared to white infants despite gestational age at delivery |
| Bullinger, L., Simon, K., and Tucker Edmonds, B. | 2022 | United States | Georgia did not expand Medicaid. While Medicaid expansion has varying effects on women based on childbearing stage, substantial gains by women of all stages occurs with expansion |
| Broussard D.L., Sappenfield W.B., Fussman C., et al | 2011 | United States | The proposed core indicators are a set of measures that all states can use to evaluate their preconception health efforts. Furthermore, the indicators serve as a basis for improving the surveillance of the health of reproductive age women. Focusing on preconception health as a means for improving pregnancy outcomes and reducing infant mortality. |
| Boulet, S., Johnson, K., Parker, C., et al. | 2006 | United States | At Grady Hospital in Atlanta, GA, 22 African American women who birthed low birth weight infants were enrolled in a pilot program where they received health care visits every 1/3 months for 2 years to address risk factors for delivering low birth weight infants. 1/4 of these women were affected by unrecognized or poorly managed chronic health problems and non wanted to become pregnant during the next two years. |
| Barreto, T., Li, C., Yoon-Kyung, C., et al. | 2021 | United States | Georgia was 1 of 9 states with an SMMI rate between 1/14 and 1/49 (the lowest category of SMMI rates). |
| Barnett, C. | 2007 | Scotland specific | Midwives should discuss the advantages of actively joining the health promotion hospitals and health promoting health service networks with their service managers.  Health promotion needs to be put firmly on the maternity map.  Key Point  Health promotion is a core quality issue and should be incorporated into the daily work of the midwife. |
| Alzate M., M. | 2006 | Georgia specific | Higher infant mortality rate between black and white population attributed to: ...less prenatal care during the first trimester (Georgia Department of Human Resources, Division of Public Health, 1997). Additionally, unlike in most developed and many developing countries, there is no mandatory vacation or paid maternity leave in the US (Albelda, 2001). ...black women’s tendency to receive less prenatal care does not explain the gap in maternal mortality between black and white women...an important factor that accounts for that gap may be differences in the quality of prenatal or obstetric care received.  This situation makes poor and low-income women in Georgia, and Georgia welfare recipients (who are mainly black) more vulnerable,and puts them at greater risk of morbidity. |
| Adams E. K., Johnston E. M. | 2016 American Journal of Public Health | United States | The Affordable Care Act provided new opportunities for coverage of women of re-productive age by expanding access to health insurance nationally and improving the quality of care covered by that insurance. However, the paper reports that as of January 2016 19 states refused to expand Medicaid. In nonexpanding states, an estimated 1.7 million women will not be eligible for either Medicaid or subsidized private policies and will still face unstable coverage during a pregnancy. |
| Anon | 2012 | Scotland specific | The Smoking, Health and Social Care (Scotland) Act 2005 has been shown to reduce the number of preterm births in Scotland |
